# Interim results of the safety and immune-efficacy of 1 versus 2 doses of COVID-19 vaccine BNT162b2 for cancer patients in the context of the UK vaccine priority guidelines

**DOI:** 10.1101/2021.03.17.21253131

**Authors:** Leticia Monin-Aldama, Adam G. Laing, Miguel Muñoz-Ruiz, Duncan R McKenzie, Irene del Molino del Barrio, Thanussuyah Alaguthurai, Clara Domingo-Vila, Thomas S. Hayday, Carl Graham, Jeffrey Seow, Sultan Abdul-Jawad, Shraddha Kamdar, Elizabeth Harvey-Jones, Rosalind Graham, Jack Cooper, Muhammad Khan, Jennifer Vidler, Helen Kakkassery, Sinha Shubhankar, Richard Davis, Liane Dupont, Isaac Francos Quijorna, Puay Lee, Josephine Eum, Maria Conde Poole, Magdalene Joseph, Daniel Davies, Yin Wu, Ana Montes, Mark Harries, Anne Rigg, James Spicer, Michael H Malim, Paul Fields, Piers Patten, Francesca Di Rosa, Sophie Papa, Tim Tree, Katie Doores, Adrian C. Hayday, Sheeba Irshad

## Abstract

**Background:** The efficacy and safety profile of vaccines against severe acute respiratory syndrome coronavirus 2 (SARS-CoV-2) have not been definitively established in immunocompromised patient populations. Patients with a known cancer diagnosis were hitherto excluded from trials of the vaccines currently in clinical use.

**Methods:** This study presents data on the safety and immune efficacy of the BNT162b2 (Pfizer-BioNTech) vaccine in 54 healthy controls and 151 mostly elderly patients with solid and haematological malignancies, respectively, and compares results for patients who were boosted with BNT162b2 at 3 weeks versus those who were not. Immune efficacy was measured as antibody seroconversion, T cell responses, and neutralisation of SARS-CoV-2 Wuhan strain and of a variant of concern (VOC) (B.1.1.7). We also collected safety data for the BNT162b2 vaccine up to 5 weeks following first dose.

**Findings:** The vaccine was largely well tolerated. However, in contrast to its very high performance in healthy controls (>90% efficacious), immune efficacy of a single inoculum in solid cancer patients was strikingly low (below 40%) and very low in haematological cancer patients (below 15%). Of note, efficacy in solid cancer patients was greatly and rapidly increased by boosting at 21-days (95% within 2 weeks of boost). Too few haematological cancer patients were boosted for clear conclusions to be drawn.

**Conclusions:** Delayed boosting potentially leaves most solid and haematological cancer patients wholly or partially unprotected, with implications for their own health; their environment and the evolution of VOC strains. Prompt boosting of solid cancer patients quickly overcomes the poor efficacy of the primary inoculum in solid cancer patients.

**RESEARCH IN CONTEXT:** *Evidence before this study:* Some cancer patients have been shown to exhibit sustained immune dysregulation, inefficient seroconversion and prolonged viral shedding as a consequence of severe acute respiratory syndrome coronavirus 2 (SARS-CoV-2) infection. Consequently, their exclusion and, in particular, the exclusion of patients receiving systemic anti-cancer therapies, from the registry trials of the 5 approved COVID-19 vaccines raises questions about the efficacy and safety of SARS-CoV-2 vaccination in this patient population. In addition, whilst the change in the UK’s dosing interval to 12-weeks aimed to maximise population coverage, it is unclear whether this strategy is appropriate for cancer patients and those on systemic anti-cancer therapies.

*Added value of this study:* We report that the RNA-based SARS-CoV-2 BNT162b2 vaccine administered in cancer patients was well tolerated, and we provide first insights into both antibody and T cell responses to the vaccine in an immunocompromised patient population.

*Implications of all the available evidence:* In cancer patients, one dose of 30ug of BNT162b2 yields poor vaccine efficacy, as measured by seroconversion rates, viral neutralisation capacity and T cell responses, at 3- and 5-weeks following the first inoculum. Patients with solid cancers exhibited a significantly greater response following a booster at 21-days. These data support prioritisation of cancer patients for an early (21-day) second dose of the BNT162b2 vaccine. Given the globally poor responses to vaccination in patients with haematological cancers, post-vaccination serological testing, creation of herd immunity around these patients using a strategy of ‘ring vaccination’, and careful follow-up should be prioritised.

## Introduction

There has been much concern about the impact of severe acute respiratory syndrome coronavirus 2 (SARS-CoV-2) exposure on groups considered at high risk of developing COVID-19, including those who suffer from cancer, many of whom are elderly, and in whom immunocompetence may be jeopardised by the malignancy and/or its treatment.^1–3^ Public health measures such as the use of masks, social distancing, test and trace programmes have helped limit virus transmission but have been insufficient to curb the pandemic.^4–7^ Thus, there was an urgent need for safe and effective prophylactic vaccines. On the 2^nd^ December 2020, COVID-19 mRNA Vaccine BNT162b2 produced by Pfizer-BioNTech was authorised by Medicines & Healthcare products Regulatory Agency (MHRA) in the UK for active immunisation to prevent COVID-19 caused by SARS-CoV-2 virus, in individuals 16 years of age and older (https://www.gov.uk/government/news/uk-authorises-pfizer-biontech-covid-19-vaccine). The active substance of the COVID-19 mRNA Vaccine BNT162b2 is a lipid-nanoparticle (LNP)– nucleoside-modified mRNA vaccine encoding full-length, uncleaved and stabilized SARS-CoV-2 Spike.^8^

The phase 3 trial for the BNT162b2 vaccine given at Day-1 followed by Day-21 booster resulted in a 95% efficacy at preventing COVID-19 illness, including severe disease.^8^ Of the 18,860 individuals immunized with the vaccine, none with an active oncological diagnosis was included. Exclusion criteria for the trial included a medical history of COVID-19, treatment with immunosuppressive therapy, or diagnosis with an immunocompromising condition. On 30^th^ December 2020, the UK announced that the second doses of the COVID-19 vaccines should be given towards the end of 12 weeks rather than in the previously recommended 3-4 weeks for all vaccinee groups. A subsequent note of caution about possible low vaccine response in immunosuppressed patients appeared in the UK government Green Book published in 12^th^ February 2021; recommending where possible to schedule the 2^nd^ dose earlier.^9^

We recently published^10^ that for some cancer patients, especially those with B cell malignancies, SARS-CoV-2 infection may result in delayed or negligible seroconversion, prolonged shedding, and sustained immune-dysregulation, as compared to the general population^11^. Likewise, some data report the suboptimal vaccine efficacy in older^12^ and in immunocompromised populations^13–15^, both of which descriptors may apply to cancer patients prioritised for early vaccination. The prospect that cancer patients might be wholly or partially unprotected by SARS-CoV-2 vaccination has implications for their individual health and for the control of virus transmission local to their environments that can include frequent visits to health-care facilities.^10,16,17^ In relation to these issues, we established the SOAP-02 vaccine study (Sars-CoV-2 fOr cAncer Patients) aimed to answer: 1) the safety and efficacy of BNT162b2 COVID-19 vaccine in cancer patients; and 2) whether or not there was a profound impact for cancer patients of boosting at day-21 post initial vaccination.

## METHODS

### Trial design, objectives and participants

We conducted a prospective longitudinal cohort study of cancer patients and healthy controls (HC), most of whom were health care workers between December 8^th^, 2020, and February 18^th^, 2021. Patients with a known diagnosis of cancer presenting at Guy’s & St Thomas’ Trust Hospital, King’s College Hospital or Princess Royal University Hospital, who were eligible for the COVID-19 mRNA BNT162b2 vaccine were screened and approached for informed consented into the SOAP-study. To facilitate cross-study comparisons we included a cohort of vaccinated HC as internal experimental controls for inter-study variability. The trial was approved by the institutional review boards of the participating institutions (IRAS ID: 282337 REC ID: 20/HRA/2031). Blood samples were captured at: pre-vaccination (time-point 1 (TP1)), at week 3 after first vaccine dose (timepoint 2 (TP2)), at week 5 after first vaccine dose (time-point 3 (TP3)). Follow-up is planned for further blood sampling after the delayed vaccine boost. Where possible serial nasopharyngeal SARS-CoV-2 rRT-PCR swab-tests were performed every 10-days or in cases of symptomatic COVID-19 (Figure 1A). The interim results of safety and efficacy for 205 participants up to 5-weeks following the first vaccination dose are reported here. Participants vaccinated between December 8^th^ to December 29^th^ received two 30-μg doses of BNT162b2, administered intramuscularly 21 days apart; but in line with changes in the national government guidelines, patients vaccinated after this date received only one dose within the study period with a planned follow up booster at 12-weeks.

**Figure 1:**
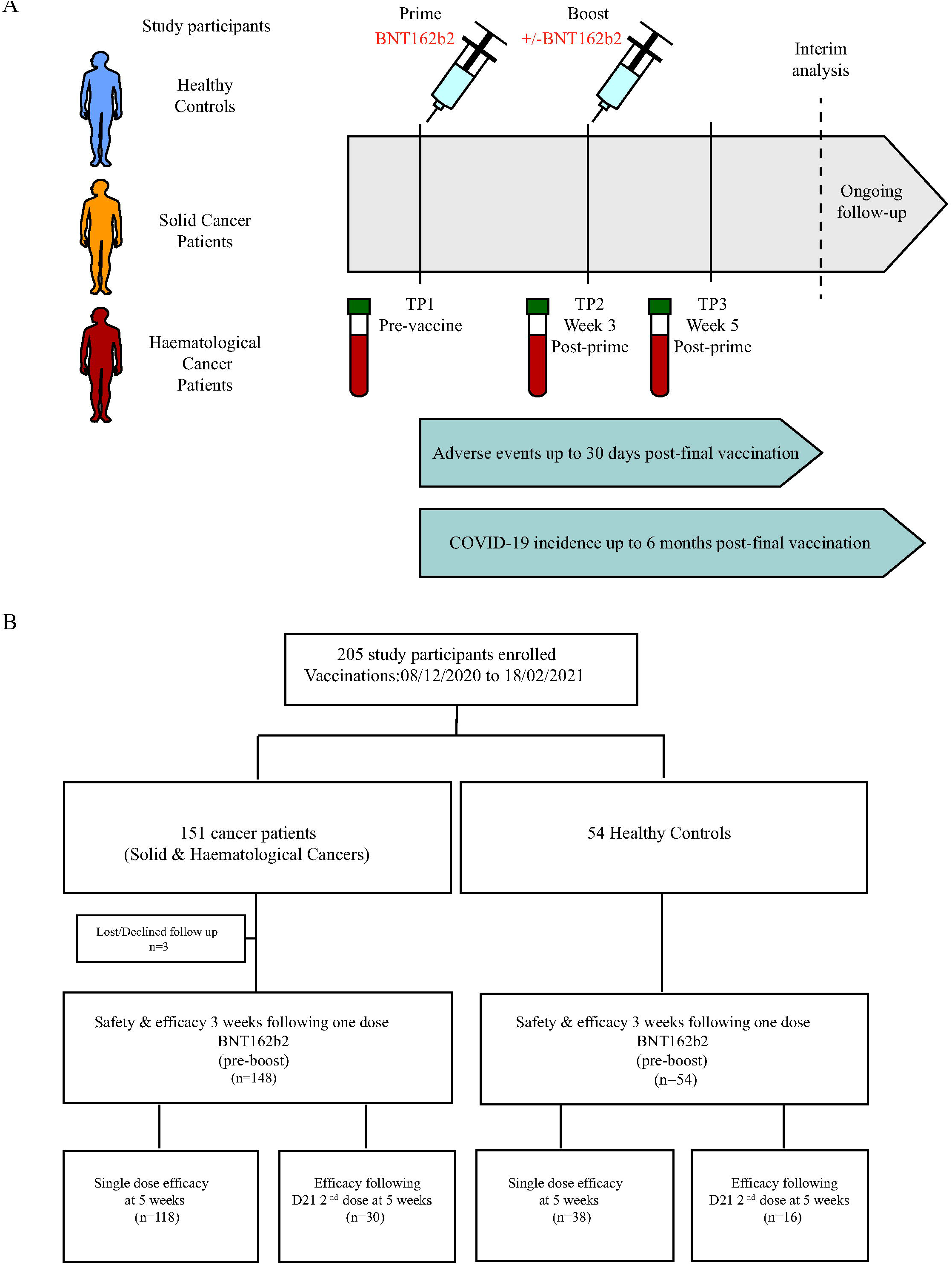
Trial design and enrolment. **A**. Schematic to show the time points at which trial activities (blood tests, nasal swab samples and safety checks) were performed in relation to the vaccination doses: Time point 1 = pre vaccine, time point 2 = 3 weeks following 1^st^ dose and time point 3 = 5 weeks following 1^st^ dose **B**. The diagram represents all enrolled participants upto 18^th^ February 2021.

### Trial end-points

The primary end-point was the humoral and cellular immunity to SARS-CoV-2 Spike (S) protein in cancer patients following the first injection of the COVID-19 mRNA BNT162b2 Vaccine. The secondary end-point of the study was the safety following each vaccine dose. Telephone consultations to evaluate reactogenicity and safety were scheduled weekly where possible. Adverse events were graded according to the following scale: mild, does not interfere with activity; moderate, interferes with activity; severe, prevents daily activity; and grade 4, emergency department visit or hospitalization. Safety data are included up to the cut-off date of February 18^th^, 2021.

### Laboratory analyses

The laboratory analyses performed on blood samples received are detailed in supplementary materials.

### Statistical analysis

Statistics were computed in R using statistical tests outlined in the figure legends. Briefly, patients between cancer groups were compared with a Kruskal-Wallis test with Dunn’s post-hoc test. Analysis of virus neutralization and patient trajectories following boosting was conducted with a paired Wilcoxon test. Immune cell counts between responder groups were compared with a Wilcoxon test. Correlations were analysed by Spearman’s test. Where appropriate, p-values were adjusted by the Benjamini-Hochberg correction to reflect multiple comparisons. Chi-squared tests were used to assess the association of serological response with cancer status.

## RESULTS

### Participants

From December 8^th^, 2020 until February 18^th^, 2021, 151 cancer patients and 54 HC consented to enrolment into the SOAP-vaccine study. Of the cancer patients, 31 cancer patients were vaccinated with BNT162b2 on days 1 and 21; and 118 primed have a planned second dose boost at around 12-weeks (Figure 1B). Of the 54 HC, 16 received vaccinations 21-days apart; and 38 primed are planned for a delayed 12-week boost. Analysis of samples and data obtained after February 18^th^, 2021 are ongoing.

The clinical characteristics of trial participants are provided (Table 1). Among cancer patients, 52% were male, 82% were white, and 64% had at least one other coexisting comorbidity. The median age was 73 years, and 75% of the participants were older than 65 years of age. Of the 151 cancer patients, 63% (95/151) represented a diversity of solid malignancies and 37% (56/151) presented with haematological malignancies (Table 1). Most solid cancer patients had late-stage disease (80%) and with a cancer diagnosis for >24months. The HC cohort comprised vaccine-eligible individuals, primarily health care workers, that did not compose an age-matched control cohort for the cancer patients, but facilitated comparison of vaccine efficacy and safety in our study with results of other studies of BTN162b2.^8,18,19^

The distribution of the anti-cancer treatments given in relation to the date of vaccine administrations for solid and haematological cancers is shown in Table 2. For solid cancers, 41.3% (38/92) of the patients had had an anti-cancer treatment within 15-days prior to the 1^st^ dose of the vaccine. Of these 38 patients, 19 had had chemotherapy, immune checkpoint inhibitors (CPI) or both. 54% (50/92) went on to have anti-cancer treatment within 15-days after administration of the first dose of the vaccine; with 50% (25/47) receiving chemotherapy, CPI or both in this timeframe. Of the 25 solid cancer patients who received the second day-21 boost, 9 (36%) had received anti-cancer treatment in the preceding 15 days; and 15 (60%) went on to have treatment in the following 15 days (Table 2). For haematological cancers, 47% (26/55) of the patients had had an anti-cancer treatment within 15-days prior to the first dose of the vaccine. Of these 26 patients, 50% had received combination therapies of either chemotherapy or targeted therapies, all with the addition of monoclonal antibodies (mAb) and 31% received targeted therapies alone. The mAb therapies were anti-CD20, anti-CD30 or anti-CD38. Targeted therapies consisted of Bruton tyrosine kinase inhibitor (BTKi), Bcl-2 inhibitor (Bcl2i) or bortezomib. 49% (27/55) received anti-cancer treatment within 15-days after the administration of the first dose of the vaccine; with 41% (11/27) receiving combination therapies in this timeframe. 8 patients were on a BTKi within 15-days before and after the vaccine administration. Only 6 haematological cancer patients received the second day-21 booster, with 2 on a BTKi at the time of vaccine administration (Table 2). Blood analyses following the second day-21 booster were conducted on 5 of these 6 patients (see below).

### Safety and tolerability

#### RNA-based SARS-CoV-2 BNT162b2 vaccine well tolerated in cancer patients

Toxicity data was available for 180 and 47 participants following the first inoculum and the second day-21 boost, respectively. 54.3% cancer and 37.5% of HC BNT162b2 recipients, reported no toxicity following the first inoculum (Figure 2Ai,ii). However, following the second day-21 boost, 71.0% of cancer patients reported no toxicity as compared to 31.25% of HC (Figure 2Aiii,iv). Additionally, only 6.8% of cancer patients reported local and systemic effects as compared to 50% of HC following the day-21 boost (Figure 2Aiii,iv). One cancer patient previously prescribed immune checkpoint inhibitors presented with deranged liver function tests requiring hospital admission 3 weeks following the first inoculum (grade 4); the cause remains unclear at this time. Routine clinical laboratory values or laboratory abnormalities observed on all other participants after BNT162b2 vaccinations at the study time-points did not demonstrate any significant findings (Supplementary Table 1). Pain at the injection site within 7 days after injection of the first dose of the vaccine was the most commonly reported local reaction (Figure 2Bi,ii). We document a noticeably lower incidence of participants reporting moderate symptoms in the cancer cohort as compared to the HCs following the first dose and second dose of the vaccine (Figure 2B,C). No differences in safety profiles between haematological and solid cancer patients were observed. Safety monitoring will continue for 18months after administration of the second dose of vaccine.

**Figure 2:**
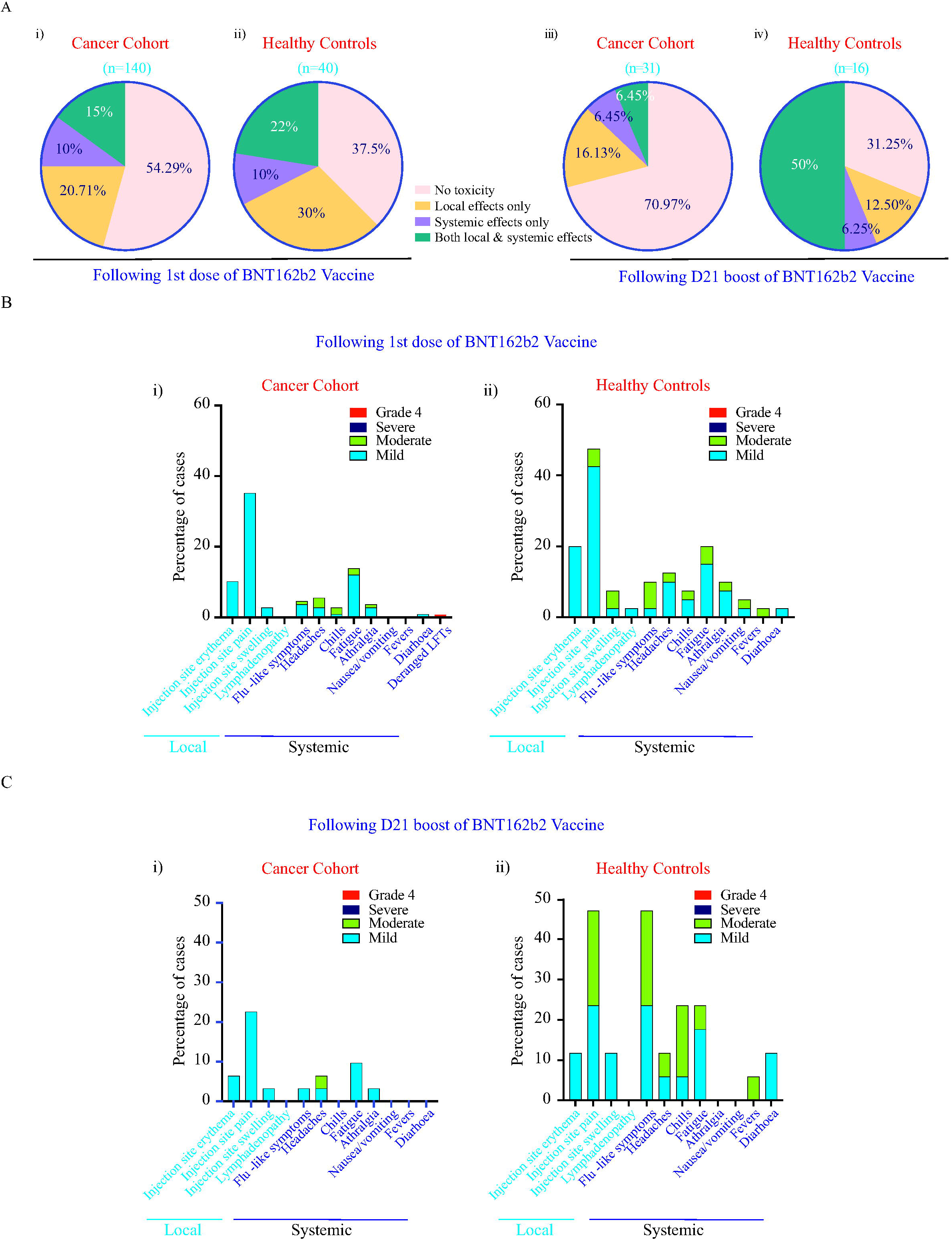
Local and systemic effects reported within 30 Days after injection of BNT162b2 in cancer patients and HCs. Data on local and systemic reactions were collected with telephone consultations with participants for 30 days after vaccination. **A**. Percentage of patients reporting no toxicity or toxicity (local effects only vs systemic effect only vs both local and systemic effects) in cancer and HCs following **i)** 1^st^ dose and **ii)** 2^nd^ Day-21 booster of BNT162b2 vaccine is shown. **B**. Breakdown of specific local and systemic side effects in cancer and HCs following 1^st^ dose **C**. Breakdown of specific local and systemic side effects in cancer and HCs following 2^nd^ day-21 booster of BNT162b2 vaccine is shown. Symptoms were assessed according to the following scale: mild, does not interfere with activity; moderate, interferes with activity; severe, prevents daily activity; and grade 4, emergency department visit or hospitalization.

### Efficacy

#### Poor efficacy of a priming vaccine inoculum in cancer patients

In assessing the immuno-efficacy of BNT162b2, it was clearly essential to exclude individuals whose immune systems may have been stimulated by past or concurrent infection. Hence, to test for asymptomatic SARS-CoV-2 infections following vaccination, cancer patient participants were asked to provide nose and throat swabs, ideally every 10-days. However, given the high prevalence of the B.1.1.7 (Kent) variant in the UK^20^, coupled with a national lockdown announced on January 4^th^ 2021 (https://www.gov.uk/government/news/prime-minister-announces-national-lockdown), only 79 patients from the study were able to attend for screening for the whole study period, of whom only twelve had more than one swab performed. Up until day-21 post-first vaccine inoculum, six positive cases were observed (Figure Supp 1A,B): two developed severe COVID-19 and succumbed to the disease, one prior to TP2 sampling; four experienced asymptomatic/mild disease, of whom two continued to swab positive at day 28 and day 36. Conversely, no new positive swab tests were recorded after 21 days post-first vaccine inoculum (Figure Supp 1A). For the reasons considered above, the 4 surviving swab-test-positive individuals were excluded from the main cohort analysis of vaccine efficacy, but are reconsidered later in this manuscript.

Antibody seroconversion was the primary assay for vaccine efficacy. As described^21^, we deployed an ELISA test to measure IgG antibodies specific for the SARS-CoV-2 S protein that was included in the vaccine. The test was validated for batch effects using standard positive and negative controls, in relation to which the threshold for a positive score was set at 70 EC50 dilution units (Figure Supp 1C). Baseline scores of >70 provided an additional means to identify individuals experiencing past or concurrent SARS-CoV-2 exposure, whose classification was further aided by our use of an ELISA assay for baseline and/or developing IgG reactivity to SARS-CoV-2 N protein that was not included in the vaccine. This ELISA too has been described^21,22^, and was re-validated (Figure Supp 1D). By these combined means, we identified 15 individuals (5 HC; 7 solid cancer patients; 3 haematological cancer patients) as possibly pre-exposed, two of whom were confirmed by virus swab test (above). Thus, 17 individuals in total were removed from the cohort analysis of vaccine efficacy but are reconsidered later in this manuscript.

When the remaining individuals were examined for anti-S IgG titres at ∼21 days following a single vaccine inoculum, they revealed significantly different levels of vaccine efficacy across the three cohorts (Table 3): thus, ∼97% (31/32) HCs were “responders” (R), whereas only ∼39% (21/54) of solid cancer patients were responders (p<0.0001), and only ∼13% (5/39) of haematological cancer patients (p<0.0001). Amongst the responders, anti-S IgG titres varied across a >100-fold range in each cohort (Figure 3A), across which the median titres were comparable in each cohort. In short, the main differences between HC, solid cancer patients, and haematological cancer patients was in the capacity to make a response rather than the scale of the response made. The HC cohort was chosen to illustrate the efficiency and scale of seroconversion that may be achieved, and for that and above mentioned reasons was not designed as a matched, cancer-free control cohort for the patients who were mostly much older. Nonetheless, the failure to make an overt response to the first vaccine inoculum did not obviously correlate with age (Figure 3B) and more likely relates to disease and treatment, as considered later.

**Figure 3.**
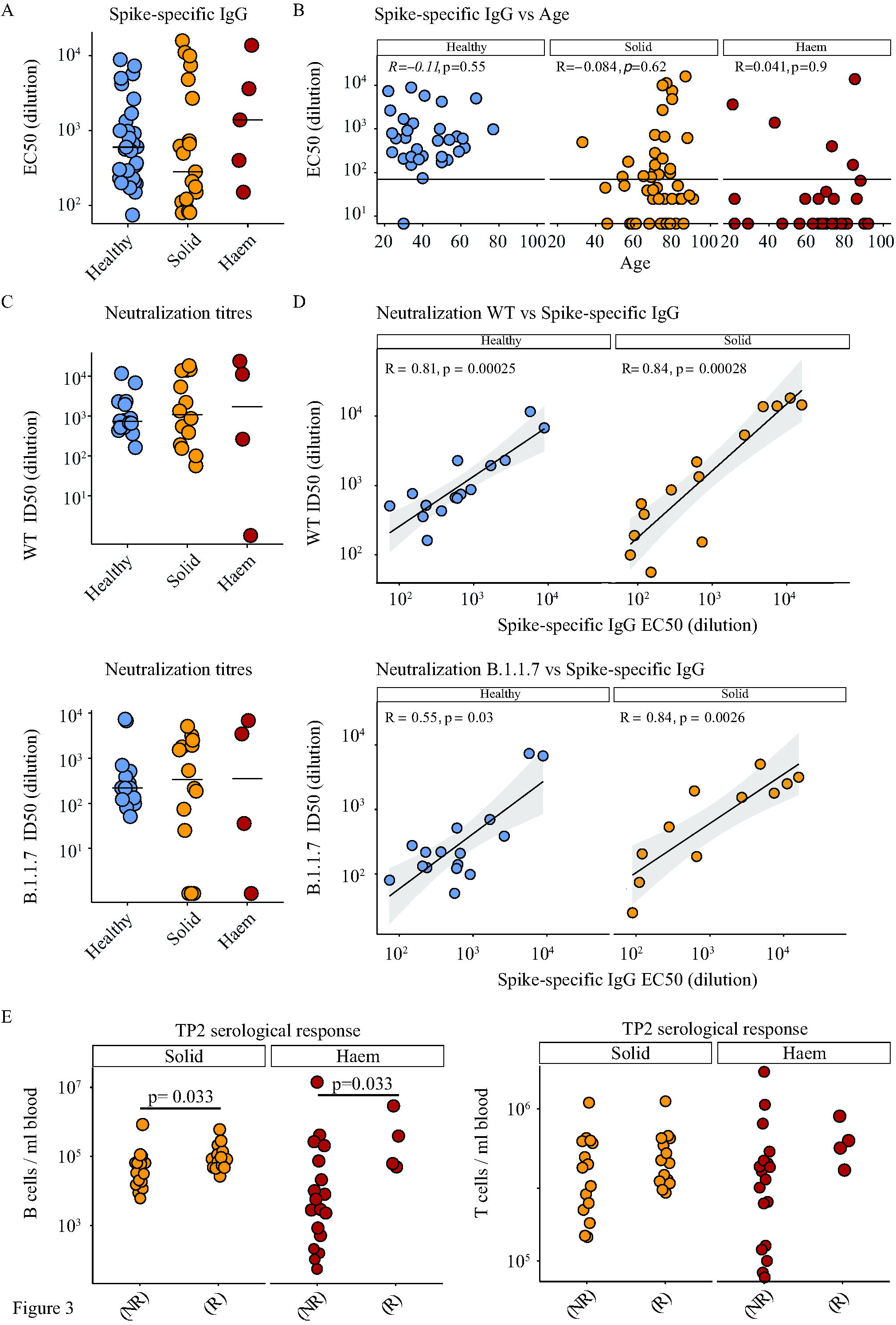
Serological response to COVID-19 vaccine BNT162b2. **A**. Spike-specific IgG titres (EC50) in plasma samples at 3 weeks post vaccine in healthy controls n=31 (blue), solid tumour cancer patients n=21 (yellow) or haematological cancer patients n=5 (red). Kruskal-Wallis test with Dunn’s post-hoc test. **B**. Association of age with serological response (Spike-specific IgG ELISA) at 3 weeks post vaccine (Spearman correlation) in healthy controls n=32 (blue), solid tumour cancer patients n=54 (yellow) or haematological cancer patients n=39 (red). **C**. Neutralization titres against WT SARS-CoV-2 and B.1.1.7 variant in plasma samples at 3weeks post vaccine in healthy controls n=16 (blue), solid tumour cancer patients n=14 (yellow) and haematological cancer patients n=4 (red). Kruskal-Wallis test with Dunn’s post-hoc test. **D**. Correlation between Spike-specific IgG titres and neutralisation titres against WT SARS-CoV2 samples at 3 weeks post vaccine in healthy controls n=16 (blue) and solid tumour cancer patients n=14 (yellow). **E**. Baseline T-cell and B-cell counts/ml of blood in non-responders (NR) and responders (R) segregated by cancer type; solid (yellow) n=46 (R=14/NR=15) and Haematological (red) n=32 (R=4/NR=18). Wilcoxon test.

To examine the functional impact of seroconversion, we examined serological responders for the capacity to neutralise infection by either the vaccine matched SARS-CoV-2 Spike (Wuhan strain, referred to as wild-type (WT), or as England 2020/02/407073, a SARS-CoV-2 strain that was pre-eminent in the UK during most of 2020), or the SARS-CoV-2 strain B.1.1.7 (Spike variant VOC 202012/01, lineage B.1.1.7) that rapidly became pre-eminent in the UK following its emergence in late-2020.^20^ To this end, we employed a recently described, validated assay^21–23^, and found that all HC responders (as judged by ID50 dilution; i.e. high values equate to potent neutralisation) had neutralising capacity although the scores were significantly lower for the B.1.1.7 strain *versus* England 02/2020/407073 (p=0.001) (Figure 3C; Supp Figure 1E). Most but not all solid cancer and haematological cancer patient responders also showed neutralisation capacity: the average ID50 value was comparable across all three cohorts, but the range was greater in the cancer patients (Figure 3C). When directly cross-compared for responders, there was a very strong correlation of anti-S IgG titres with neutralisation scores for HC and solid cancer patients, there being too few haematological cancer patient responders to justify the comparison (Figure 3D).

In sum, a priming inoculum of 30μg of BNT162b2 induced seroconversion in the great majority of HCs, but failed to achieve this in most solid and haematological cancer patients, with efficacy in the haematological cancer cohort being significantly worse than in solid cancer patients. Moreover, not all seroconverted cancer patients showed virus neutralisation. By comparing outcomes with baseline flow cytometry data, it became clear, unsurprisingly, that non-responder (NR) status correlated significantly with reduced B cell numbers in solid cancer patients (median value for no-responders = 34,680 cells/ml *versus* 87,840 cells/ml for responders) and in haematological cancer patients (median value for non-responders= 5,040 cells/ml *versus* 225,020 cells/ml for responders) (Figure 3E). There was likewise a trend by which NR status correlated with reduced peripheral blood T cell numbers (Figure 3E).

#### T cell responses to a priming vaccine inoculum

From a subset of individuals in each cohort we subjected PBMC to fluorospot assays to quantitate T cells secreting IFN-γ and/or IL-2 in response to stimulation with two separate SARS-CoV-2 spike protein peptide pools, one spanning the S2 domain and one spanning the Receptor Binding Domain (RBD), which is not contained within S2.^18,24^ Peptides within these pools are capable of stimulating both MHCI- and MHCII-restricted T cell responses.^24,25^ Additionally, we compared responses to peptides from commonly encountered viruses, CMV, EBV, and influenza and from tetanus that were contained in two pools, CEFT and CEF that likewise favoured stimulation of MHCII-restricted and MHCI-restricted T cells, respectively. The assay was stringently assessed for batch effects by use of a documented positive control and a validated negative control (Figure Supp 2A,B). A single cut-off for T cell responses was determined by receiver operated characteristic (ROC) analysis using pooled IFN-γ and IL-2 and responses to RBD and S2 from infection-naïve HC individuals before and 21 days following priming inoculum, establishing a threshold value of >7 cytokine secreting cells/10^6^ PBMC as a positive response (Figure Supp 2A,B).

Of HCs assayed, 14 of 17 (82%) showed T cell responses at day-21 post-vaccination, manifesting in IFN-γ and/or IL-2 production, and many examples of double-producing cells (Figure 4A; Figure Supp 2C; Table 3). These responses correlated with serological responses, as shown by bi-variate analyses, although three serological responders failed to show T cell reactivity while there was one case of T cell reactivity in a HC non-responder (Figure 4B). The situation for solid cancer patients was different by two notable and related criteria. First, 22 of 31 patients (∼71%) showed T cell responses (Figure 4A), an efficacy much more comparable to HC than was true for seroconversion (above). Second, whereas only 1 serological responder failed to show T cell reactivities, T cell reactivities were evident for 8 serological non-responders (Figure 4B). Although the dynamic range of positive values was broad (from 0 to >300), the median value for patients was somewhat lower than for HCs (7 *versus* 25 for S2-IFN-γ reactivity; Figure 4A).

**Figure 4.**
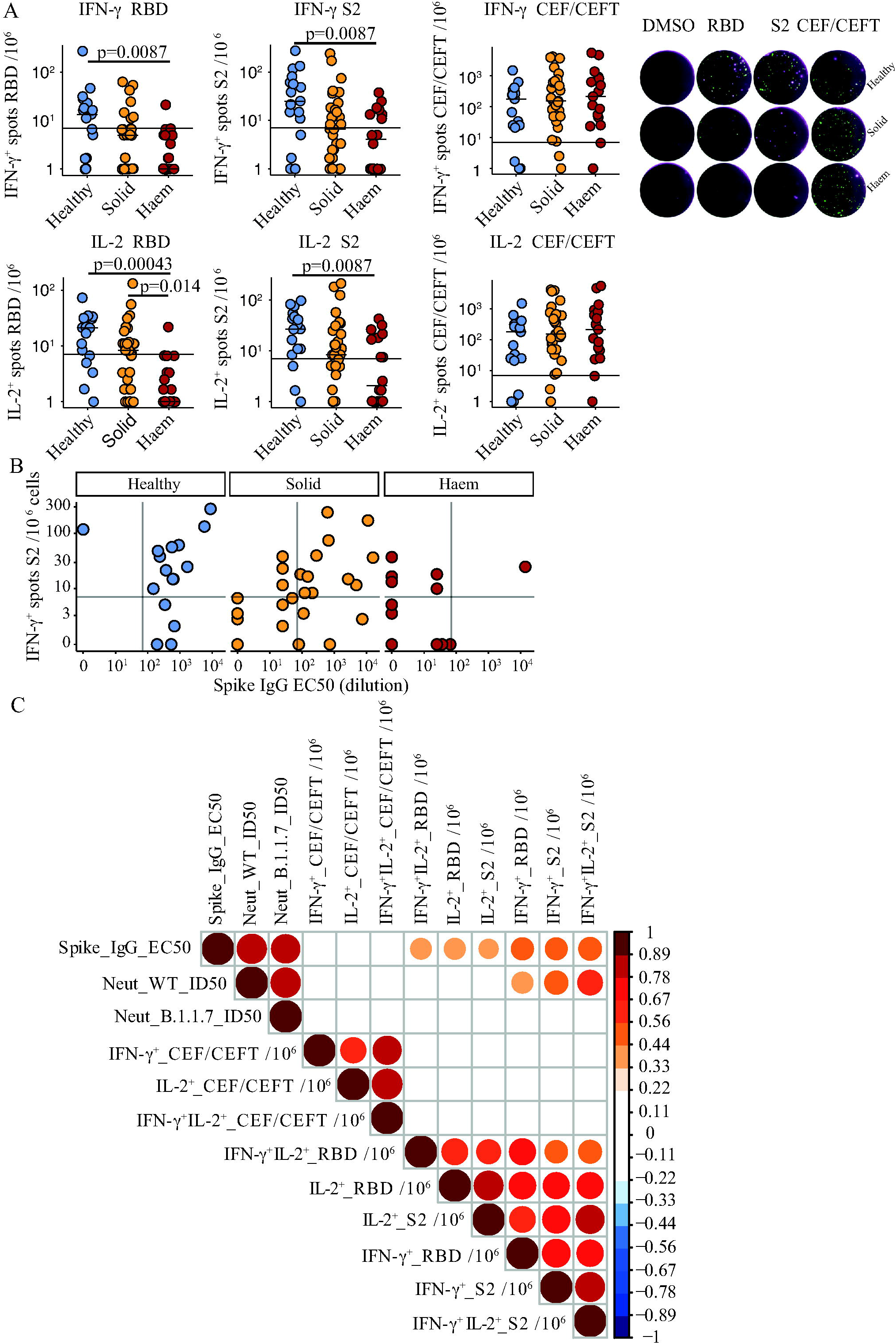
T-cell response to COVID-19 vaccine BNT162b2. **A**. Left: IFN-γ^+^ and IL-2^+^ response to stimulation with peptides from RBD, S2, and CEF-CEFT reported as number of spots per 10^6^ cells in PBMC samples at 3 weeks post vaccine in healthy controls n=17 (blue), solid tumour cancer patients n=31 (yellow) or haematological cancer patients n=18 (red). Bar represents median value by group, horizontal line represents threshold of specific response. Kruskal-Wallis test with Dunn’s post-hoc test. Right: Exemplary Fluorospot images for PBMC taken at TP2 (21d after 1^st^ inoculation). IFN-γ-secreting cells are shown in green, IL-2 secreting cells in magenta and cells co-secreting both cytokines are visualised as white. **B**. Relationship between serological response and T-cell response in healthy control n=16 (blue), solid tumour cancer patients n=31 (yellow) and haematological cancer patients n=18 (red). horizontal and vertical lines represent threshold of specific response for T-cell and serological response respectively. **C**. Spearman correlation between T-cell responses (fluorospot counts/10^6^ PBMC) and serological responses as determined by ELISA and neutralization assay. Colour scale indicates spearman R-value, all p values <0.01.

This phenomenon was largely echoed by haematological cancer patients. Thus, of 18 assayed for T cell reactivities at day-21, 9 (50%) scored positive for IFN-γ and/or IL-2 in response to S2 peptides (Figure 4a). Although the median value for S2-reactive IFN-γ-secreting cells was comparable to that of solid cancer patients, the dynamic range was less (from 0 to 37.5; Figure 4A). Importantly, this was not because T cell response competence *per se* was compromised in haematological cancer patients because the frequencies and strengths of CEF+CEFT responses were quantitatively comparable across all cohorts (Figure 4A). Interestingly, 8 of 9 haematological cancer patients showing T cell responses were serological non-responders (Figure 4A, B). Moreover, positivity for RBD peptides was rare. Given that RBD-reactivity is possibly more specific for SARS-CoV-2 than is reactivity to S2, which sequence is highly conserved with minor variation in common cold coronaviruses, it is possible that seronegative haematological cancer patients showing low-level T cell reactivities reflect pre-existing T cell reactivities rather than those induced by the vaccine.

In sum, a priming inoculum of 30μg of BNT162b2 induced T cell responses to S2 and RBD in the majority of HCs and solid cancer patients, although many of the cancer patients were serological non-responders. When viewed across all subjects, IFN-γ production in response to S2 or RBD correlated with neutralisation of England 2020/02/407073 but did not correlate as strongly as anti-Spike IgG (Figure 4C). Moreover, T cell responses did not correlate significantly with B.1.1.7 neutralisation (Figure 4C).

#### Primed versus boosted immunity

The next step was to investigate whether the subsequent trajectories of primary responses might be positively affected by boosting with 30μg of BNT162b2. Thus, the cohorts were divided into two subsets, boosted or not at day-21, and then were compared at TP3, which was 5 weeks after the priming inoculum and 14 days after the boost for those who received it. There was some attrition in sampling at TP3 relative to TP2, particularly for those who were not boosted. Nonetheless, the results were striking by several criteria. First, 100% (12/12) of boosted HC scored positive (Table 3), including the seroconversion of 1 individual who was formally seronegative (above). Second, 18 of 19 (95%) solid cancer patients scored as seropositive after boost (Table 3), including the seroconversion of 8 individuals, whereas only 9 of 21 (43%) who were not boosted scored as seropositive (Table 3), comparable to a seropositivity rate of 39% for solid cancer patients attained at week 3 with priming inoculum alone (above). Third, individual trajectory plots (Figure 5A) clearly showed that without boosting, TP3 anti-Spike IgG titres remained mostly comparable to TP2 in HC and in solid cancer patients, with some tendency to decline. Conversely, the boost substantially increased anti-S IgG titres, significantly in solid cancer patients (p<0.03) (Figure 5A), such that their average was now comparable to that of HCs, notably representing a much younger cohort.

**Figure 5.**
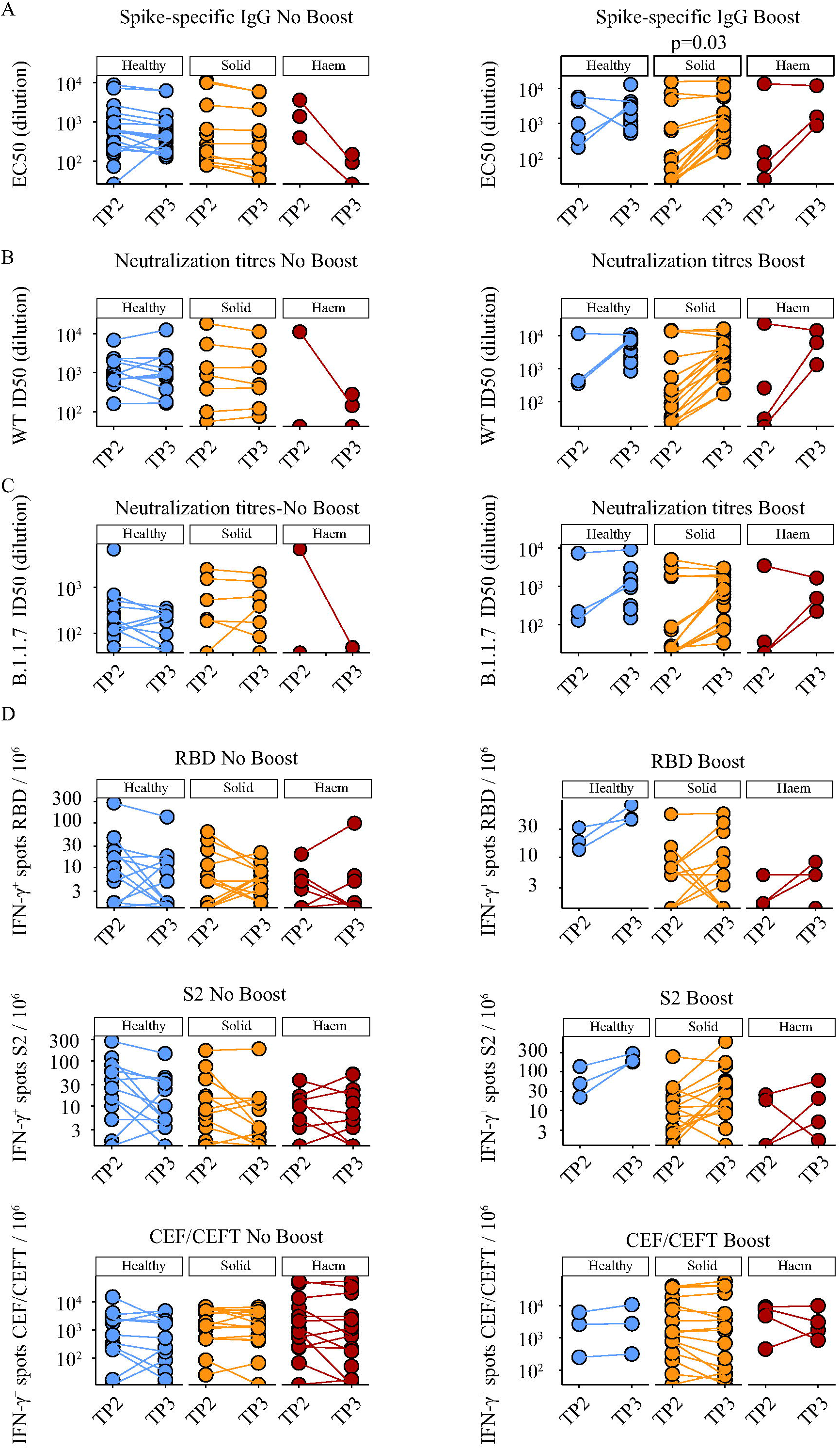
Comparison of single-dose vs prime/boost with COVID-19 vaccine BNT162b2. **A**. Spike-specific IgG titres in plasma samples at 3 and 5 weeks post vaccine in individuals receiving a single vaccine dose (No Booster); healthy controls n=26 (TP2) n=18 (TP3) (blue), solid tumour cancer patients n=14 (TP2) n=12 (TP3) (yellow), haematological cancer patients n=3 (TP2/3) (red), and in individuals receiving two doses (Booster); healthy controls n=6 (TP2) n=12 (TP3) (blue), solid tumour cancer patients n=15 (TP2) n=18 (TP3) (yellow) or haematological cancer patients n=4 (TP2) n=3 (TP3) (red). **B**. Neutralization titres against WT SARS-CoV2 in plasma samples at 3 and 5 weeks post vaccine in receiving a single vaccine dose (No Booster); healthy controls n=14 (TP2) n=12 (TP3) (blue), solid tumour cancer patients n=7 (TP2/3) (yellow), haematological cancer patients n=2 (TP2) n=3 (TP3) (red), and in individuals receiving two doses (Booster); healthy controls n=3 (TP2) n=11 (TP3) (blue), solid tumour cancer patients n=15 (TP2) n=18 (TP3) (yellow) or haematological cancer patients n=4 (TP2) n=3 (TP3) (red). **C**. Neutralization titres against B.1.1.7 SARS-CoV2 in plasma samples at 3 and 5 weeks post vaccine individuals receiving a single vaccine dose (No Booster); healthy controls n=14 (TP2) n=11 (TP3) (blue), solid tumour cancer patients n=7 (TP2/3) (yellow), haematological cancer patients n=2 (TP2) n=3 (TP3) (red), and in individuals receiving two doses (Booster); healthy controls n=3 (TP2) n=11 (TP3) (blue), solid tumour cancer patients n=15 (TP2) n=18 (TP3) (yellow) or haematological cancer patients n=4 (TP2) n=3 (TP3) (red). **D**. Cytokine response to stimulation with peptides from RBD, S2, and CEF-CEFT reported as number of spots per 10^6^ PBMC at 3 and 5weeks post vaccine in individuals receiving a single vaccine dose (No Booster) healthy controls n=14 (TP2) n=13 (TP3) (blue), solid tumour cancer patients n=15 (TP2/3) (yellow) or haematological cancer patients n=14 (TP2) n=18 (TP3) (red) and in individuals receiving two doses (Booster); healthy controls n=3 (TP2/3) (blue), solid tumour cancer patients n=16 (TP2/3) (yellow) or haematological cancer patients n=4 (TP2/3) (red). a-d) Paired Wilcoxon test.

For haematological cancer patients, it was not possible to assess rigorously the impact of boosting because only 6 were eligible for the booster before government guidelines mandated a longer delay post priming. Nonetheless, of those 6 who were boosted, analyses were performed on 5, of whom 3 scored positive (Table 3). Moreover, of 25 analysed at TP3 without prior boosting, only 2 scored positive and there were some conspicuous reductions in anti-S IgG titres between TP2 and TP3 (Figure 5A).

For solid cancers, the considerable impact of the boost on anti-S IgG titres was mirrored by profound increases in the capacity to neutralise both Wuhan strain and B.1.1.7 SARS-CoV-2 Spike proteins assayed (Figure 5B,C). Boosting likewise increased virus neutralisation by HC sera, and this increase was particularly evident for B.1.1.7, the current major circulating strain in the UK (Figure 5B,C). Again, few haematological cancer patients showed neutralisation capacity, and there was considerable heterogeneity among them with respect to neutralisation potency (Figure 5B,C).

A substantial quantitative impact of boosting was apparent for T cells in HC and cancer patients. Thus, of 3 HCs, 4 haematological cancer patients, and 16 solid cancer patients that were all boosted, only two solid cancer patients and one haematological cancer patients did not display SARS-CoV-2-peptide reactive IFN-γ and/or IL-2 T cell responses (Figure 5D). Moreover, boosting induced the acquisition of T cell responses in 1 haematological cancer patient and 4 solid cancer patients. By contrast, among those HCs and solid cancer patients who were not boosted, there was acquisition of T cell reactivity at TP3 in only two HCs who scored negative at TP2 (Table 3), and TP2-existing T cell responses either remained comparable at TP3 or declined somewhat, e.g. IL-2 and IFN-γ responses to RBD peptides. (Figure 5D). As a control, the values for CEF and CEFT cell responses remained mostly the same from TP2 to TP3, irrespective of boosting (Figure 5D). Again, few haematological cancer patients were boosted, but for those who were the potency of the T cell response was increased (Figure 5D). Nonetheless, there was no case of acquired T cell reactivity at TP3 among those who were negative at TP2 whether or not they were boosted (Table 3).

In sum, by three weeks following a primary inoculum of BNT162b2, cancer patients in general showed poor efficacy in terms of seroconversion and virus neutralisation, and over the following two weeks, there was essentially no increase in responders’ IgG or neutralisation capacity and no increase in the numbers seroconverting. Moreover, whereas a primary inoculum was somewhat better at inducing T cell than antibody responses, T cell reactivity too did not improve naturally over the following two weeks, showing some tendency to wane. By contrast, each of those metrics was significantly increased in solid cancer patients by boosting at day-21.

#### Impacts of disease, treatments, and pre-exposure

Given that most cancer patients did not seroconvert following a primary vaccine inoculum, we investigated in greater detail factors that might associate with poor responsiveness, scrutinising types of malignancy, treatment, and baseline immunophenotypes, albeit that the heterogeneity of the vaccine cohorts undermined statistical power in several areas. This notwithstanding, we found that serological non-responders, which were the most common phenotype among haematological cancers, were spread evenly across patients with B-, T-, and myeloid-cell malignancies (Figure 6A). For solid cancers, non-responders were again spread comparably across diverse tumour types, with some enrichment in respiratory tract cancers (Figure 6B). Non-responders were also somewhat more frequent among those who received the vaccine within 15 days of cancer treatment, particularly chemotherapy (Figure 6C). Indeed, 8/9 patients who were serological non-responders but T cell responders had had intensive treatments, including those depleting B cells, before or after the vaccine inoculum. This notwithstanding, the very high effectiveness of boosting in solid cancer patients suggested its power to counteract factors mitigating vaccine efficacy.

**Figure 6.**
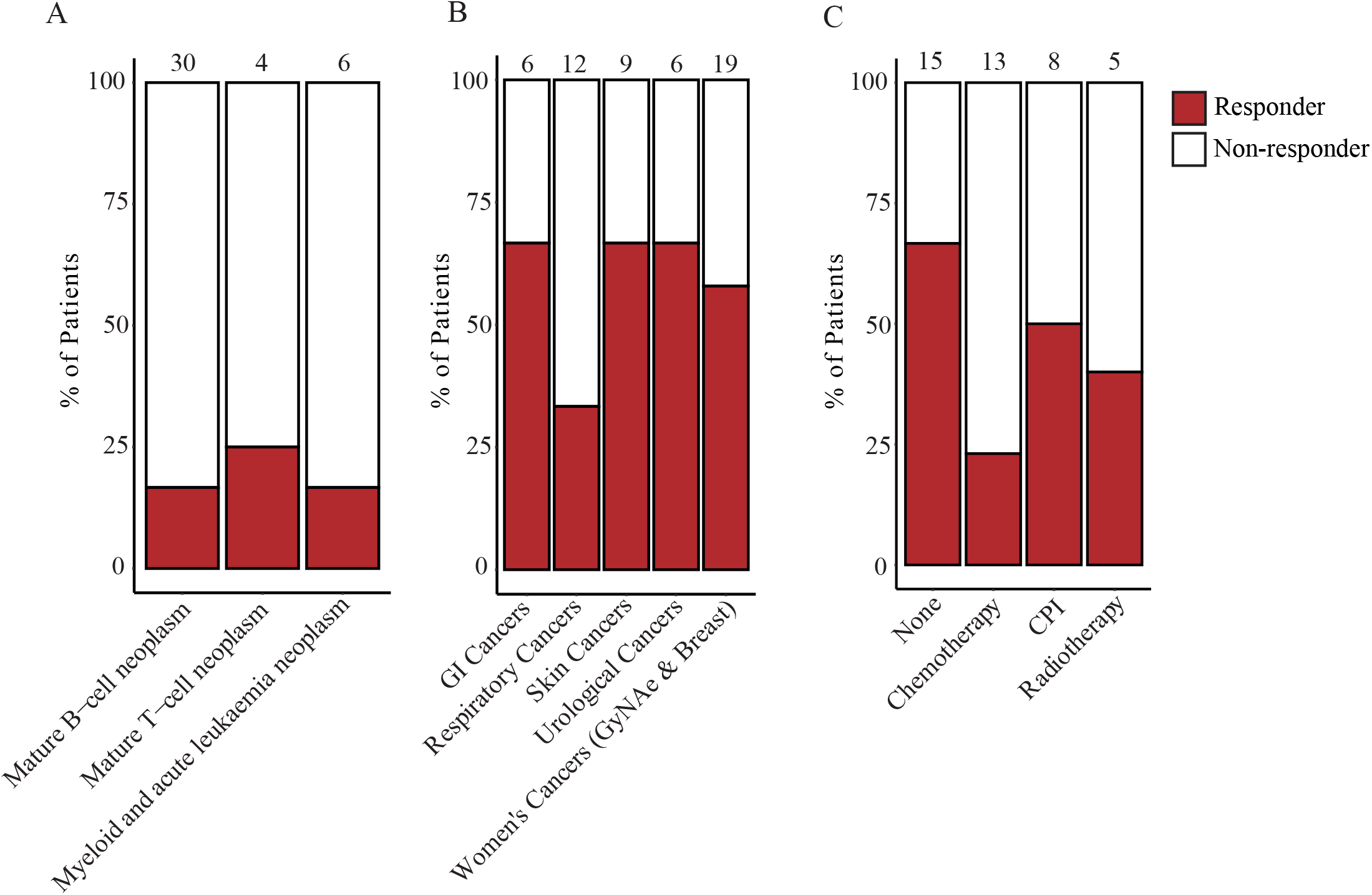
Impact of disease and treatments on serological response. **A**. The number of serological responders and non-responders were assessed across haematological cancer patients at TP2 (n=40), and segregated by the type of cancer. **B**. The number of serological responders and non-responders were assessed across solid cancer patients at TP2 (n=52), and segregated by the type of cancer. **C**. The number of serological responders and non-responders were assessed across solid cancer patients at TP2 (n=41), and segregated based on the type of treatment received within 15 days either side of first vaccination.

As considered above, 17 individuals were excluded from the main study comparisons because of suspected SARS-CoV-2 exposure. Nonetheless, when their serological and T cell responses were viewed, they showed considerable heterogeneity across the dynamic ranges described for the main cohorts (Figure Supp 3). Thus, swab-test and/or serological evidence indicative of prior or concurrent exposure did not guarantee stronger vaccine responses in the small number of study participants examined here.

## DISCUSSION

Here, we present the first report of safety and immune efficacy (a.k.a. immunogenicity) for any COVID-19 vaccine in immunocompromised patient populations, specifically, those with an active cancer diagnosis. The RNA-based SARS-CoV-2 BNT162b2 (Pfizer-BioNTech) vaccine administered in cancer patients was generally well tolerated, even in those on immunotherapy where exacerbated immune activity might have been deemed a risk. Nonetheless, single dose vaccination was profoundly less immunogenic in cancer patients compared to healthy controls (HCs). Notably, anti-SARS-CoV-2 IgG responses at week 3 following the first dose were 39% and 13% in the solid and haematological cancers, compared to 97% in HCs. Immunological parameters in cancer patients did not improve in the subsequent two weeks without boosting. Crucially, however, each immunological metric measured significantly and substantially improved in solid cancer patients following a day-21 boost; to date, numbers of boosted haematological cancer patients are insufficient to assess its impact.

A two-dose regimen of BNT162b2 (30μg per dose, given 21 days apart) was reported to be 95% effective against Covid-19 in non-immunocompromised patient populations.^8^ Patients with immunocompromise due to solid tumour and haematological cancers, organ transplantation, haematopoietic stem cell transplantation, and autoimmune diseases on biological agents were excluded from COVID-19 vaccine clinical trials. Previous studies on seasonal vaccines have reported suboptimal vaccine efficacy in immunocompromised populations.^13–15^ Our data show that following first dose, most solid and haematological cancer patients remained wholly or partially immunologically unprotected up until at least 5 weeks following primary inoculum. Thus, delaying the day-21 boost for these patients may have serious implications, both for the individual and for controlling the pandemic. As we and others have reported, immunocompromised patients have a higher incidence of harbouring persistent SARS-CoV-2 infections^10,16,17^, possibly representing an important reservoir for the emergence of novel viral variants.^26,27^ This may likewise apply to any patients who might reasonably be assumed to be immunocompromised. From these perspectives, the case is made for urgently re-evaluating UK policy for the BNT162b2 dosing interval for all cancer patients, and other high-risk groups, *prima facie* consistent with the JCVI’s update (12^th^ February 2021) recognising that specific populations may mount inferior responses.^9^ Additionally, studies to investigate the immunogenicity of more frequent boosts in immunocompromised patients who remain seronegative are warranted.

Although correlates of protection against COVID-19 remain incompletely defined, it is broadly considered that neutralizing antibodies and antigen-specific T cells will be required for vaccine efficacy.^28,29^ Our immunogenicity results after a single BNT162b2 vaccine dose showed that >90% of HC generated cytokine-producing T cells and/or anti-SARS-CoV-2 IgG, and displayed virus-neutralizing capacity, in agreement with other efficacy data.^19^ Nonetheless, this does not necessarily mean that vaccine boosting has negligible impact in HCs, since parameters such as durable immunological memory were not measured. Moreover, it is striking that a single vaccine inoculum was not as effective at inducing neutralisation of VOC.B.1.1.7 *versus* the WT strain, but that this was improved in several participants by boosting. This seems germane to our concerns about the potential for VOCs to emerge under the umbrella of partial protection.

A single vaccine inoculum evidently induced IFN-γ and/or IL-2-producing RBD-reactive and S2-reactive T cells in solid cancer patients, but the proportion of seroconversion in this cohort was less than half than that of HC, with consequent deficiencies in virus neutralization. In contrast, 95% of solid cancer patients who received the BNT162b2 vaccine according to the approved day-21 prime-boost regimen seroconverted 2 weeks after boost, and their antibodies neutralized both the Wuhan and the B.1.1.7 strain. Hence, a policy of day-21 boosting rapidly conferred immunological protection on a hitherto largely unprotected population. Given the incidence of solid cancers, this amounts to a very large population.

Patients with haematological malignancies are reportedly at increased risk of adverse outcomes from SARS-CoV-2^30^ and given this vulnerability, there is an urgent need to protect this population expeditiously. Thus, the extremely poor immunogenicity following a single vaccine inoculation of haematological cancer patients is of particular concern. Clearly, this is not an entirely unexpected finding given the immunobiology of the underlying diseases and the fact that many patients with mature B cell malignancies have experienced prior or concurrent B cell depleting agents. Nonetheless, although our interim analysis was insufficiently powered to assess the impact of the day-21 boost in these patients, it is strikingly clear that increased measures are urgently required to confer immunological protection, most likely comprising prompt vaccine boosting and frequent serological monitoring. Until such measures are introduced, it seems important that this population continues to observe all COVID-19-associated measures such as social distancing and shielding when attending hospitals, even after vaccination. Moreover, whereas cancer patients in the UK were assigned vaccination priority level 4, no prioritisation was given to non-professional carers and/or immediate social contacts, who may potentially transmit virus to incompletely protected patients and/or be infected by them. Possibly these groups should be considered as priorities in future pandemic planning, in part to limit nodes of increased transmission and VOC emergence.

It might be argued that irrespective of immunogenicity data, evidence for protection is provided by there being no new positive swab tests post day-21. Moreover, it has been considered that SARS-CoV-2 infection may *de facto* provide a boost to incompletely protected patients. We consider these viewpoints as currently unsubstantiated, noting that 17 individuals suspected of prior exposure did not all make strong responses to the vaccine, and that several haematological patients with S2-specific T cell reactivity did not acquire T cell reactivity to RBD. Thus, we conclude that no understanding was available of the risk of changing from a planned day-21 boost to a delayed 12-week boost in cancer patients at the time of UK policy change. Our BNT162b2 immunogenicity data raises serious concerns that maximising population coverage for the general population with the first dose has come at a price of increased risk to “clinically extremely vulnerable” groups, for whom we suggest an urgent review of current UK vaccine strategy.

## Data Availability

Upon request

## Acknowledgements

We thank patients and blood donors consenting in this study and the medical and research teams at Guy’s and St. Thomas Trust (GSTT) hospitals. We thank members of the GSTT and King’s College Hospital (KCH) trial teams who contributed to patient recruitment for the SOAP study at GSTT and KCH hospitals; and clinical colleagues at GSTT, KCH, and PRUH for assisting with patient identification and sample collection. The SOAP study (IRAS 282337) is sponsored by King’s College London and GSTT Foundation NHS Trust. It is funded from grants from the KCL Charity funds to S.I. (PS10822), Cancer Research UK to S.I. (C56773 / A24869), program grants from Breast Cancer Now including S.I. at King’s College London and to the Breast Cancer Now Toby Robin’s Research Center at the Institute of Cancer Research, London. This work was also supported by the Wellcome Trust Investigator Award to A.C.H. (106292/Z/14/Z), the Rosetrees and John Black Charitable Foundation award to A.C.H (11130), the Cancer Research UK Cancer Immunotherapy Accelerator and a UK COVID-Immunology-Consortium (CIC) grant to AH (C33499/A20265); the King’s Together Rapid COVID-19 Call awards to K.J.D and M.H.M and A.C.H; the Foundation Dormeur, Vaduz for funding equipment to K.J.D; the Huo Family Foundation Award to M.H.M and K.J.D; the Cancer Research Institute Irvington Fellowship (D.R.M); the MRC-KCL Doctoral Training Partnership in Biomedical Sciences (C.G., MR/N013700/1); and the Francis Crick Institute (A.C.H.), which receives core funding from Cancer Research UK (FC001093), the MRC (FC001093) and the Wellcome Trust (FC001093).

## Author contributions

Conceptualization and study design (AGL, IdMdB, TA, TSH, AH, SI); Investigation (LM, AGL, MMR, DRM, IdMdB, CDV, TSH, CG, JS, SAJ, HK, SS, RG, RD, LD, SK, IFQ, PL, JE, MCP, RG, MJ, DD, YW, MHM); Patient recruitment (TA, EHJ, JC, MK, AM, MH, JS, PF, PP); Clinical database (TA, EHJ, JC, MK); Formal analysis (LM, AGL, MMR, DRM, IdMdB, TSH, SK); Data curation (IdMdB); Methodology (AGL, IdMdB); Visualization (LM, AGL, MMR, DRM, IdMdB, CDV, TT, SI); Writing – original draft (AH, SI); Writing – review & editing (AH, SI, FdiR, AM, MH, JS, PF, PP, TT, KD); Funding acquisition (AH, SI); Supervision (AH, SI, TT, KD).

## Figure legends

**Supplementary figure 1. Natural infection and neutralisation responses following BNT162b2 vaccination**.

**A**. Incidence of PCR swab-confirmed SARS-CoV-2 infection within patients and healthy controls following first dose of vaccine.

**B**. Progression of PCR swab-confirmed SARS-CoV-2 infection following first dose of vaccine.

**C**. Spike-specific IgG titres prior to vaccination in healthy controls (n=14), solid tumour cancer patients (n=59) and haematological cancer patients (n=34), illustrating seropositivity threshold (horizontal line at EC50 dilution of 70) and suspected pre-exposed individuals.

**D**. N-specific IgG titres prior to and following vaccination, highlighting PCR-swab confirmed and baseline Spike-high healthy controls (n=3) and solid tumour cancer patients (n=5) (red), and other healthy controls (n=2), solid tumour cancer patients (n=4) and haematological cancer patients (n=3) suspected of pre-existing or developing SARS-CoV-2 infection due to high N IgG titres (grey). Horizontal line indicates N-specific IgG threshold for suspected infection. White dots indicate remaining healthy controls (n=41), solid tumour cancer patients (n=60) and haematological cancer patients (n=42) from combined vaccination cohorts.

**E**. Neutralization titres against WT and B.1.1.7 strains SARS-CoV-2 from sera of healthy controls (n=16) at 3 weeks post-vaccination. Paired Wilcoxon test.

**Supplementary figure 2. QC and combined T-cell responses to COVID-19 vaccine BNT162b2**

**A**. IFN-γ^+^ and IL-2^+^ response to stimulation with peptides from RBD and S2 was reported as number of spots per 10^6^ cells in a negative and a positive PBMC sample that was run alongside for every assay. Variation in the response among different batches was assessed with CVs as follows: IFN-γ in S2 (1.702, 0.198), IFN-γ in RBD (1.380, 0.155), IL-2 in S2 (0.943, 0.222), IL-2 in RBD (1.133, 0.209) for the negative and the positive controls, respectively.

**B**. Determining a cut-off value for assigning positive and negative fluorospot responses. Graph represents a ROC plot showing assay diagnostic sensitivity against specificity (one minus proportion of false positives) following detection of IFN-γ and IL-2 fluorospot responses to RBD and S2 peptide pools in healthy control subjects before and following one dose of BNT162b2. By convention, we selected the cut-off value that provides an operating position nearest to that of the “perfect test” (i.e., closest approximation to 100% sensitivity and 100% specificity), which was ≥7 cytokine secreting cells/10^6^ PBMC.

**C**. IFN-γ ^+^ and IL-2^+^ dual producers after stimulation with peptides from RBD, S2 and CEF-CEFT was reported as number of spots per 10^6^ cells in PBMC samples at 3weeks post vaccine in healthy controls n=17 (blue), solid tumour cancer patients n=31 (yellow) or haematological cancer patients n=18 (red). Bar represents median value by group, horizontal line represents T cell response threshold (7). Kruskal-Wallis test with Dunn’s post-hoc test.

**Supplementary figure 3: Serological and T cell responses in vaccinated individuals with concomitant SARS-CoV-2 infection**.

Patients returning a positive SARS-CoV-2 PCR swab and those displaying baseline seropositivity to Spike or high N-specific IgG titres at any point (all red; PCR swab positive also diamond) were excluded from the prior analysis. Here they are shown in the context of other patients who were not suspected of natural infection (white). Lines link repeated samples from individual patients.

**A**. Spike-specific IgG titres in suspected naturally infected healthy controls (n=5), solid tumour cancer patients (n=9), haematological cancer patients (n=3), as compared to the remaining cohort of healthy controls (n=41), solid tumour cancer patients (n=60) and haematological cancer patients (n=42). Horizontal line indicates Spike seropositivity threshold (70).

**B**. IFN-γ+ T cell response to S2 peptides in suspected naturally infected healthy controls (n=3), solid tumour cancer patients (n=4), haematological cancer patients (n=1), as compared to the remaining cohort of healthy controls (n=17), solid tumour cancer patients (n=31) and haematological cancer patients (n=22). Horizontal line indicates T cell response threshold (7).

**C**. IL-2^+^ T cell response to S2 peptides in suspected naturally infected healthy controls (n=3), solid tumour cancer patients (n=4), haematological cancer patients (n=1), as compared to the remaining cohort of healthy controls (n=17), solid tumour cancer patients (n=31) and haematological cancer patients (n=22). Horizontal line indicates T cell response threshold (7).

## References

1. Liu C, Zhao Y, Okwan-Duodu D, Basho R, Cui X. COVID-19 in cancer patients: risk, clinical features, and management. Cancer Biol Med 2020; 17:519–527.

2. Vijenthira A, Gong IY, Fox TA, et al. Outcomes of patients with hematologic malignancies and COVID-19: a systematic review and meta-analysis of 3377 patients. Blood 2020; 136:2881–2892.

3. Tagliamento M, Spagnolo F, Poggio F, et al. Italian survey on managing immune checkpoint inhibitors in oncology during COVID-19 outbreak. Eur J Clin Invest 2020; 50:e13315.

4. Wilder-Smith A, Freedman DO. Isolation, quarantine, social distancing and community containment: pivotal role for old-style public health measures in the novel coronavirus (2019-nCoV) outbreak. J Travel Med 2020; 27:taaa020.

5. Shen J, Duan H, Zhang B, et al. Prevention and control of COVID-19 in public transportation: Experience from China. Environ Pollut 2020; 266:115291.

6. Matzinger P, Skinner J. Strong impact of closing schools, closing bars and wearing masks during the Covid-19 pandemic: results from a simple and revealing analysis. 2020; medRxiv 2020; doi: 10.1101/2020.09.26.20202457.

7. Chang S, Pierson E, Koh PW, et al. Mobility network models of COVID-19 explain inequities and inform reopening. Nature 2021; 589:82–87.

8. Polack FP, Thomas SJ, Kitchin N, et al. Safety and Efficacy of the BNT162b2 mRNA Covid-19 Vaccine. N Engl J Med 2020; 383:2603–2615.

9. https://assets.publishing.service.gov.uk/government/uploads/system/uploads/attachment_data/file/961287/Greenbook_chapter_14a_v7_12Feb2021.pdf

10. Abdul-Jawad S, Baù L, Alaguthurai T, et al. Acute immune signatures and their legacies in severe acute respiratory syndrome coronavirus-2 infected cancer patients. Cancer Cell 2021; 39:257–275.

11. Laing AG, Lorenc A, Del Molino Del Barrio I, et al. A dynamic COVID-19 immune signature includes associations with poor prognosis. Nat Med 2020; 26:1623–1635.

12. Collier DA, Ferreira Iatm, Datir R, et al. Age-related heterogeneity in neutralising antibody responses to SARS-CoV-2 following BNT162b2 vaccination. medRxiv 2021; doi: 10.1101/2021.02.03.21251054.

13. Lo W, Whimbey E, Elting L, Couch R, Cabanillas F, Bodey G. Antibody response to a two-dose influenza vaccine regimen in adult lymphoma patients on chemotherapy. Eur J Clin Microbiol Infect Dis 1993; 12:778–782.

14. Mazza JJ, Yale SH, Arrowood JR, et al. Efficacy of the influenza vaccine in patients with malignant lymphoma. Clin Med Res 2005; 3:214–220.

15. Nordøy T, Aaberge IS, Husebekk A, et al. Cancer patients undergoing chemotherapy show adequate serological response to vaccinations against influenza virus and Streptococcus pneumoniae. Med Oncol 2002; 19:71–78.

16. Avanzato VA, Matson MJ, Seifert SN, et al. Case Study: Prolonged Infectious SARS-CoV-2 Shedding from an Asymptomatic Immunocompromised Individual with Cancer. Cell 2020; 183:1901-1912.e9.

17. Aydillo T, Gonzalez-Reiche AS, Aslam S, et al. Shedding of Viable SARS-CoV-2 after Immunosuppressive Therapy for Cancer. N Engl J Med 2020; 383:2586–2588.

18. Sahin U, Muik A, Vogler I, et al. BNT162b2 induces SARS-CoV-2-neutralising antibodies and T cells in humans. medRxiv 2020; doi: 10.1101/2020.12.09.20245175.

19. Skowronski DM, De Serres G. Safety and Efficacy of the BNT162b2 mRNA Covid-19 Vaccine. N Engl J Med 2021, Feb 17;Online ahead of print; doi: 10.1056/NEJMc2036242.

20. Davies NG, Abbott S, Barnard RC, et al. Estimated transmissibility and impact of SARS-CoV-2 lineage B.1.1.7 in England. Science 2021, Mar 3; Online ahead of print; doi: 10.1126/science.abg3055.

21. Seow J, Graham C, Merrick B, et al. Longitudinal observation and decline of neutralizing antibody responses in the three months following SARS-CoV-2 infection in humans. Nat Microbiol 2020; 5:1598–1607.

22. Pickering S, Betancor G, Galão RP, et al. Comparative assessment of multiple COVID-19 serological technologies supports continued evaluation of point-of-care lateral flow assays in hospital and community healthcare settings. PLOS Pathogens 2020; 16:e1008817.

23. Graham C, Seow J, Huettner I, et al. Impact of the B.1.1.7 variant on neutralizing monoclonal antibodies recognizing diverse epitopes on SARS-CoV-2 Spike. bioRxiv 2021; doi 10.1101/2021.02.03.429355.

24. Sahin U, Muik A, Derhovanessian E, et al. COVID-19 vaccine BNT162b1 elicits human antibody and TH1 T cell responses. Nature 2020; 586:594–599.

25. Braun J, Loyal L, Frentsch M, et al. SARS-CoV-2-reactive T cells in healthy donors and patients with COVID-19. Nature 2020; 587:270–274.

26. Choi B, Choudhary MC, Regan J, et al. Persistence and Evolution of SARS-CoV-2 in an Immunocompromised Host. N Engl J Med 2020; 383:2291–2293.

27. Kemp SA, Collier DA, Datir RP, et al. SARS-CoV-2 evolution during treatment of chronic infection. Nature 2021, Feb 5; Online ahead of print; doi: 10.1038/s41586-021-03291-y.

28. Dagotto G, Yu J, Barouch DH. Approaches and Challenges in SARS-CoV-2 Vaccine Development. Cell Host Microbe 2020; 28:364–370.

29. Kim DS, Rowland-Jones S, Gea-Mallorquí E. Will SARS-CoV-2 Infection Elicit Long-Lasting Protective or Sterilising Immunity? Implications for Vaccine Strategies (2020). Frontiers in Immunology 2020; 11:571481.

30. Passamonti F, Cattaneo C, Arcaini L, et al. Clinical characteristics and risk factors associated with COVID-19 severity in patients with haematological malignancies in Italy: a retrospective, multicentre, cohort study. Lancet Haematol 2020; 7:e737–e745.

