## Supplementary figures and images for "Interim results of the safety and immune-efficacy of 1 versus 2 doses of COVID-19 vaccine BNT162b2 for cancer patients in the context of the UK vaccine priority guidelines"

### Supplemental Figure 1

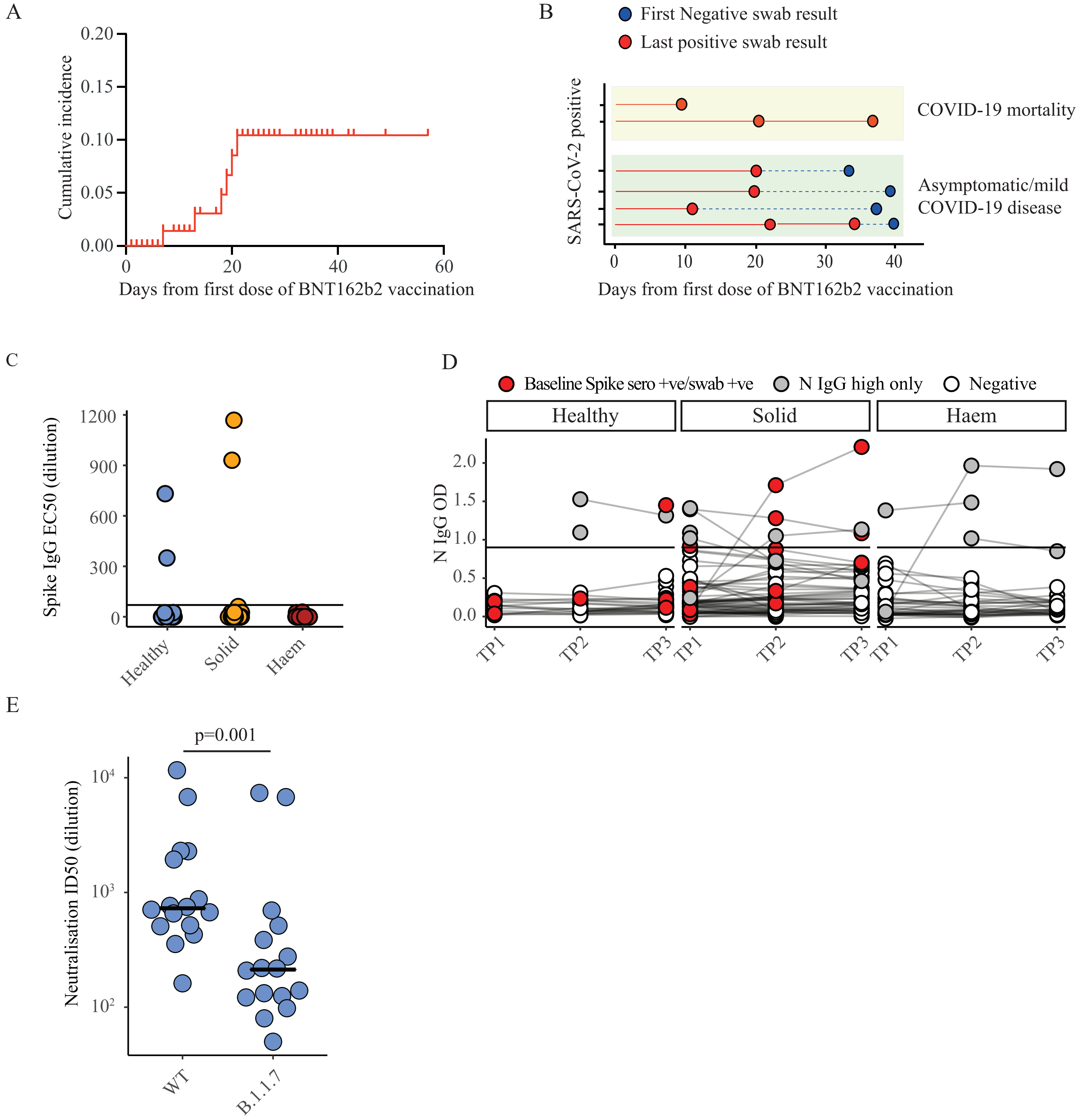


Supp Figure 1

### Supplemental Figure 2

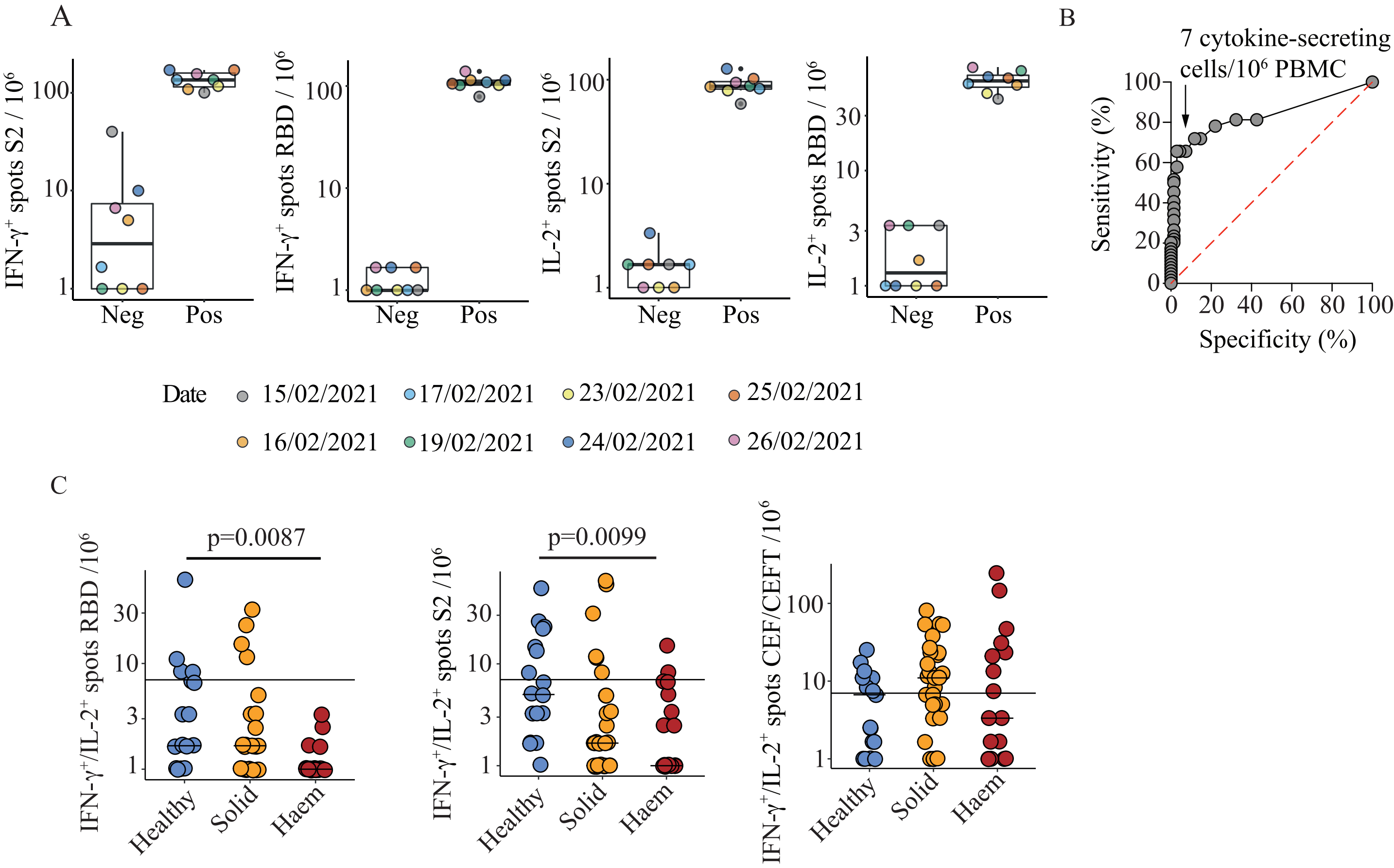


Supp Figure 2

### Supplemental Figure 3

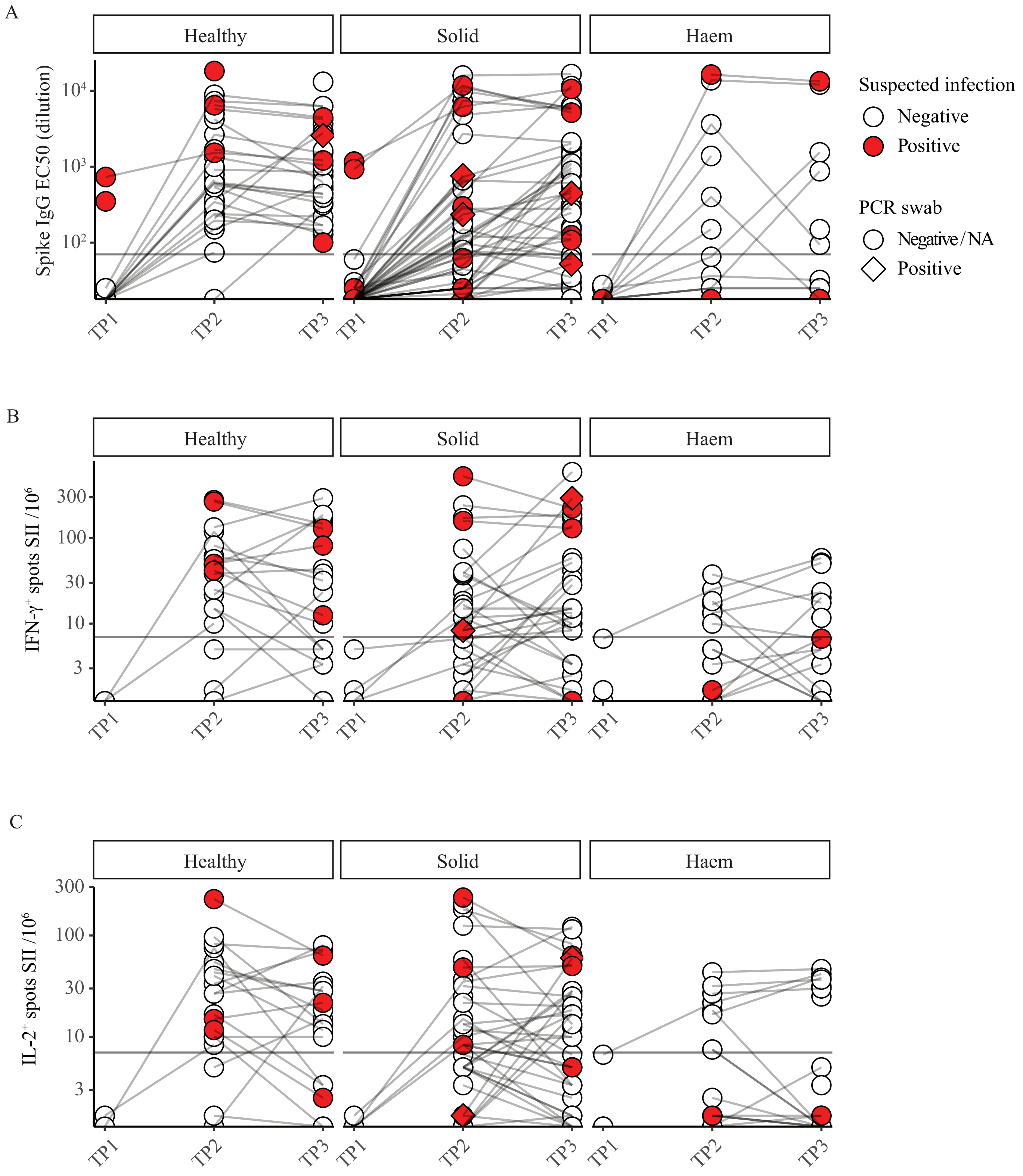
