## Supplementary material for "Interim results of the safety and immune-efficacy of 1 versus 2 doses of COVID-19 vaccine BNT162b2 for cancer patients in the context of the UK vaccine priority guidelines": Tables 1-3

**Table 1: Clinical characteristics of vaccinated cancer and healthy controls**

|  | Cancer | HC |
| --- | --- | --- |
| *n=* | *151* | *54* |
| Age |  |  |
| *Median (days)* | 73 | 40.5 |
| *Range* | 19-94 | 22-78 |
| Sex |  |  |
| *Male* | 52% (78) | 52% (28) |
| *Female* | 48% (73) | 48% (26) |
| Race |  |  |
| *Caucasian* | 82% (124) | 61% (33) |
| *BAME* | 18% (27) | 39% (21) |
| Non-oncological comorbidities |  |  |
| *Cardiovascular disease (IHD, HTN, Hypercholesteremia)* | 41% (62) | 0 |
| *Diabetes* | 15% (22) | 0 |
| *Underlying Lung Pathology* | 7.9% (12) | 0 |
| *None of above* | 36.4% (55) | 0 |
| Solid Malignancies | **95** |  |
| *Women's Cancers (Gynae, breast)* | *35% (33)* |  |
| *Urological Cancers (Renal, Prostate, testicular, bladder)* | *16% (15)* | *N/A* |
| *Skin Cancers (Melanoma, Merkel cell)* | *13% (12)* |  |
| *Thoracic malignancies (Lung, Mesothelioma)* | *22% (21)* |  |
| *GI Cancers (Stomach, oesophageal, pancreas, CRC)* | *13% (12)* |  |
| *Others* | *2% (2)* |  |
| Haematological malignancies | **56** |  |
| Mature B -cell neoplasms | 68% (38) |  |
| *Chronic lymphocytic leukaemia/Small lymphocytic lymphoma* | *11* |  |
| *Plasma Cell Myeloma* | *9* |  |
| *Diffuse large B cell lymphoma* | *8* |  |
| *Follicular lymphoma* | *4* |  |
| *Lymphoplasmacytic lymphoma* | *1* |  |
| *Burkitt’s lymphoma* | *1* |  |
| *Mantle cell Lymphoma* | *1* |  |
| *MALT lymphoma* | *1* |  |
| *Nodular sclerosing Hodgkin lymphoma* | *1* |  |
| *Post-transplant lympho-proliferative disorder* | *1* |  |
| Mature T cell neoplasms | 9% (5) |  |
| *Anaplastic large cell lymphoma* | *4* |  |
| *Angioimmunoblastic T-cell lymphoma* | *1* |  |
| Myeloid and acute leukaemia neoplasm | 18% (10) |  |
| *Acute myeloid leukaemia* | *3* |  |
| *Myelodysplastic Syndrome/Myeloproliferative Neoplasm (MDS/MPN)* | *2* |  |
| *Chronic myelomonocytic leukaemia (CMML)* | *2* |  |
| *T-cell precursor acute lymphoblastic leukaemia* | *2* |  |
| *Myelofibrosis* | *1* |  |
| Others | 5% (3) |  |
| *Osteomyelofibrosis* | *1* |  |
| *Amyloid light-chain (AL) amyloidosis* | *1* |  |
| *Erdheim-chester disease* | *1* |  |
| TNM Staging * (solids only) | *% (n)* |  |
| *1* | *8.42% (8)* |  |
| *11* | *6.30% (6)* |  |
| *111* | *27.4% (26)* |  |
| *1V* | *56.8% (54)* |  |
| *Missing data* | *1.10% (1)* |  |
| Time from cancer diagnosis to study recruitment |  |  |
| *<3months* | *23% (34)* |  |
| *3-12 months* | *20% (30)* |  |
| *12-24months* | *16% (24)* |  |
| *>24months* | *35% (53)* |  |
| *Missing data* | *7% (10)* |  |

**Table 2: Anti-cancer treatments prior or after vaccination administration**

| Solid Cancers (n=92) * | | | |
| --- | --- | --- | --- |
|  | ***0-15 days*** | ***16-29 days*** | ***>30-days*** |
| Prior to 1^st^ dose | ***41.3% (n=38)*** | ***13% (n=12)*** | ***46% (n=42)*** |
| *Treatment naïve/no treatment* | *-* | *-* | *7* |
| *Chemotherapy* | *11* | *7* | *11* |
| *Immune Checkpoint inhibition (ICI)* | *5* | *2* | *6* |
| *Chemotherapy + ICI* | *3* | *1* | *2* |
| *Targeted therapies* | *10* | *2* | *1* |
| *Endocrine therapies* | *8* | *-* | *3* |
| *Radiotherapy* | *1* | *-* | *4* |
| *Surgery* | *-* | *-* | *8* |
| Following 1^st^ dose | ***54% (n=50)*** | ***20% (n=18)*** | ***26% (n=24)*** |
| *Treatment naïve/no treatment* | *-* | *-* | *22* |
| *Chemotherapy* | *13* | *2* | *-* |
| *Immune Checkpoint inhibition (ICI)* | *8* | *2* | *2* |
| *Chemotherapy + ICI* | *4* | *1* | *-* |
| *Targeted therapies* | *10* | *3* | *-* |
| *Endocrine therapies* | *9* | *1* | *-* |
| *Radiotherapy* | *6* | *8* | *-* |
| *Surgery* | *-* | *1* | *-* |
| Prior to the Day 21 2^nd^ dose (n=25) | ***36% (n=9)*** | ***64% (n=16)*** |  |
| *Treatment naïve/no treatment* | *-* | *9* |  |
| *Chemotherapy* | *3* | *1* |  |
| *Immune Checkpoint inhibition (ICI)* | *-* | *3* |  |
| *Chemotherapy + ICI* | *-* | *1* |  |
| *Targeted therapies* | *3* | *2* |  |
| *Endocrine therapy* | *1* | *-* |  |
| *Radiotherapy* | *2* | *-* |  |
| After the Day 21 2^nd^ dose (n=25) | ***60% (n=15)*** | ***40% (n=10)*** |  |
| *Treatment naïve/no treatment* | *-* | *9* |  |
| *Chemotherapy* | *3* | *1* |  |
| *Immune Checkpoint inhibition (ICI)* | *3* | *-* |  |
| *Chemotherapy + ICI* | *1* | *-* |  |
| *Targeted therapies* | *5* | *-* |  |
| *Endocrine therapy* | *1* | *-* |  |
| *Radiotherapy* | *2* | *-* |  |
| Haematological Cancers (n=55) * | | | |
|  | ***0-15 days*** | ***16-29 days*** | ***>30-days*** |
| Prior to 1^st^ dose (n=55) | ***47% (n=26)*** | ***11% (n=6)*** | ***42% (23)*** |
| *Treatment naïve/no anti-cancer treatment* |  | *-* | *11* |
| *Chemotherapy (mini-CHOP, hydroxycarbamide, mercaptopurine, methotrexate)* | *2* | *2* | *2* |
| *Targeted therapies (e.g.BTKi, Bcl2i, bortezomib)* | *8* | *1* | *-* |
| *Single agent mAb (Anti-CD30, Anti-CD20)* | *1* | *1* | *-* |
| *Chemo/targeted therapies + immunotherapy (e.g., anti-CD20, Anti-CD30, Anti-CD38)* | *13* | *1* | *1* |
| *Lenalidomide* | *1* | *1* | *2* |
| *Immune checkpoint inhibitor* | *-* | *-* | *1* |
| *Radiotherapy* | *1* | *-* | *4* |
| *Surgery* |  | *-* | *1* |
| Following 1^st^ dose (n=55) | ***49% (27)*** | ***51% (28)*** |  |
| *Treatment naïve/no anti-cancer treatment* |  | *18* |  |
| *Chemotherapy (mini-CHOP, hydroxycarbamide, mercaptopurine, methotrexate)* | *5* | *1* |  |
| *Targeted therapies (BTKi)* | *6* | *2* |  |
| *Single agent mAb (Anti-CD30, Anti-CD20)* | *1* | *2* |  |
| *Chemo/targeted therapies + immunotherapy (e.g., anti-CD20, Anti-CD30, Anti-CD38)* | *11* | *4* |  |
| *Lenalidomide* | *3* | *-* |  |
| *Immune checkpoint inhibitor* |  | *1* |  |
| *Radiotherapy* | *1* |  |  |
| Prior to the Day 21 2^nd^ dose (n=6) | ***33% (n=2)*** | ***67%(n=4)*** |  |
| *No anti-cancer treatment* | *-* | *4* |  |
| *BTK inhibitor* | *2* |  |  |
| Following the Day 21 2^nd^ dose (n=6) | ***33% (n=2)*** | ***67%(n=4)*** |  |
| *No anti-cancer treatment* | *-* | *4* |  |
| *BTK inhibitor* | *2* |  |  |

** missing data solid cancers: n=3; haematological cancers: n=1*

**Table 3:** Efficacy of BNT162b2 vaccine

|  | **Priming inoculum efficacy at 3-weeks** | **Efficacy at 5-weeks** | |
| --- | --- | --- | --- |
|  |  | **No boost** | **day-21 boost** |
| **Anti-SARS-CoV-2 IgG response** | | | |
| HC | 97% (31/32) | 100% (18/18) | 100% (12/12) |
| Solid Cancers | 39% (21/54) | 43% (9/21) | 95% (18/19) |
| Haematological Cancers | 13% (5/39) | 8% (2/25) | 3/5* |
| **T-cell vaccine response** | | | |
| HC | 82% (14/17) | 69% (9/13) | 3/3* |
| Solid Cancers | 71% (22/31) | 53% (8/15) | 88% (14/16) |
| Haematological Cancers | 50% (9/18) | 33% (6/18) | 3/4* |

**insufficient numbers for clinical interpretation*
