## Supplemental Table 1 for "Interim results of the safety and immune-efficacy of 1 versus 2 doses of COVID-19 vaccine BNT162b2 for cancer patients in the context of the UK vaccine priority guidelines"

**Supplementary Table**

**Table 1:** Routine full blood count performed at each trial specific visit.

| Routine blood parameters | | | | |
| --- | --- | --- | --- | --- |
| Timepoint | **Pre-Vaccine** | **3-weeks following one dose of BNT162b1 vaccine** | **5-weeks following one dose of BNT162b1 vaccine** | **At 5weeks following D1 &D21 dosing** |
| Solid Cancers | | | | |
| Total numbers | 75 | 65 | 21 | 19 |
| Hb | 121 (79-175) | 119 (15-152) | 113 (93-135) | 121 (81-149) |
| WCC | 6.5 (3-19) | 6.3 (2.1-25) | 4.3 (1.1-18) | 6.3 (3.8-11.6) |
| Platelets | 245 (90-688) | 231 (195-668) | 210 (52-989) | 276 (122-411) |
| Neutrophils | 4 (1.2-16) | 4.2 (1-22.5) | 2.6 (0.7-15.3) | 4.6 (1.9-7.8) |
| Lymphocytes | 1.5 (1.2-16) | 1.4 (0.4-5) | 1.1 (0.4-3.2) | 1.5 (0.7-4.7) |
| Basophils | 0.1 (0-1.5) | 0.1 (0-0.4) | 0.1 (0-0.2) | 0.1 (0-0.2) |
| Eosinophils | 0.1 (0-0.8) | 0.1 (0-1.9) | 0.2 (0-0.5) | 0.1 (0-0.2) |
| Monocytes | 0.8 (0-1.5) | 0.6 (0.1-1.9) | 0.7 (0-1.6) | 0.6 (0.3-1) |
| CRP | 14.2 (0-78) | 4 (1-149) | 3 (1-111) | 2 (1-45) |
| Haematological Cancers | | | | |
| Total numbers | 38 | 34 | 8 | 2 |
| Hb | 127 (70-127) | 119 (74-171) | 116.5 (95-146) | N/A |
| WCC | 5.2 (1-101) | 4.6 (0.5-51.7) | 3.6 (1.1-18) |  |
| Platelets | 185 (13-473) | 166 (51-514) | 145 (52-276) |  |
| Neutrophils | 5.5 (0.1-13.1) | 2.7 (0.4-22.9) | 2.1(0.7-15.3) |  |
| Lymphocytes | 1.6 (0.2-97) | 1.05 (0.1-179) | 0.8 (0.4-3.2) |  |
| Basophils | 0.1 (0-0.9) | 0.1 (0-0.3) | 0.1 (0-0.1) |  |
| Eosinophils | 0.1 (0-0.5) | 0.1 (0-0.7) | 0.2 (0-0.3) |  |
| Monocytes | 0.8 (0.1-2.5) | 0.5 (0-2.1) | 0.5 (0-1.6) |  |
| CRP | 2 (1-144) | 3 (1-75) | 1.5 (1-15) |  |
