## Supplemental Materials for "Interim results of the safety and immune-efficacy of 1 versus 2 doses of COVID-19 vaccine BNT162b2 for cancer patients in the context of the UK vaccine priority guidelines"

**Supplementary Materials**

**LABORATORY ANALYSES**

**Sample processing and cell isolation**

Fresh whole blood samples were collected in heparin tubes (BD Biosciences) and sample processing was performed under enhanced Biosafety Level 2 conditions. Aliquots of blood from heparin tubes were stained for whole-blood flow cytometry panels (see below) or centrifuged at 2,000g for 10 min and plasma stored at −80 °C. Remaining heparinized blood was diluted with 50% volume PBS, layered over Ficoll (GE Healthcare) in Leucosep tubes (Greiner Bio-One) and centrifuged at 800g for 15 min without brake at room temperature (RT). The peripheral blood mononuclear cell (PBMC) fraction was then washed three times in cold PBS and frozen at -80C in CS10 Cryostor using MrFrosty containers before being transferred to liquid nitrogen for long-term storage.

**Flow cytometry staining and acquisition**

50μl of whole blood was stained in 50μl of antibody staining mix (**Supp Table 2**) and incubated for 20 minutes at RT before washing in Dulbecco-PBS and fixation for 10 min with Cellfix (BD Biosciences). Red blood cell lysis was performed twice using eBioscience 1x RBC lysis buffer for 15 min at room temperature, whereupon samples were spun down and resuspended in 200 μl of staining buffer for acquisition by flow cytometry. 100 μl of sample was analysed on a four-laser BD LSR Fortessa acquired with a BD high-throughput sampler, and populations were gated in FlowJo as described previously.^11^

**ELISA protocol**

ELISAs were conducted as previously described.^22^ All plasma samples were heat-inactivated at 56 °C for 30 min before use. High-binding ELISA plates (Corning, 3690) were coated with antigen (Nuclear (N) protein or the S glycoprotein at 3µg ml^−1^ (25 µl per well) in PBS, either overnight at 4 °C or for 2 h at 37 °C. Wells were washed with PBS-T (PBS with 0.05% Tween-20) and then blocked with 100 µl of 5% milk in PBS-T for 1 h at room temperature. The wells were emptied and serial dilutions of plasma (starting at 1:25, 6-fold dilution) were added and incubated for 2 h at room temperature. Control reagents included CR3009 (2 µg ml^−1^)(N-specific monoclonal antibody), CR3022 (0.2 µg ml^−1^)(S-specific monoclonal antibody), negative control plasma (1:25 dilution), positive control plasma (1:50), and blank wells. Wells were washed with PBS-T. Secondary antibody was added and incubated for 1h at room temperature. IgG was detected using goat-anti-human-Fc-AP (alkaline phosphatase) (1:1,000) (Jackson, catalogue no. 109-055-098) and wells were washed with PBS-T and AP substrate (Sigma) was added and plates read at 405 nm. ELISA measurements were performed in duplicate. EC50 values were calculated in GraphPad Prism. Where an EC50 was not reached at 1:25, a plasma was considered seropositive if the OD at 405nm was 4-fold above background^22^ and a value of 25 was assigned.

**SARS-CoV-2 (wild-type and B.1.1.7) pseudotyped virus preparation.**

Pseudotyped HIV virus incorporating the SARS-Cov-2 England 2 (England 2020/02/407073, hereafter referred to as WT) or B.1.1.7 strain Spike was prepared as previously described.^23^ Pseudotyped viruses produced in a 10 cm dish seeded the day prior with 5x106 HEK293T/17 cells in 10 mL of complete Dulbecco’s Modified Eagle’s Medium (DMEM-C, 10% foetal bovine serum (FBS) and 1% Pen/Strep (100 IU/mL penicillin and 100 mg/mL streptomycin)). Cells were transfected using 90 mg of PEI-Max (1 mg/mL, Polysciences) with: 15 µg of HIV-luciferase plasmid, 10 µg of HIV 8.91 gag/pol plasmid and 5 µg of SARS-CoV-2 Spike protein plasmid (wild-type or B.1.1.7 in pCDNA3.1). Pseudotyped virus was harvested after 72 hours, filtered through a 0.45mm filter, concentrated by ultracentrifugation and stored at -80°C until required.

**Neutralization assay with SARS-CoV-2 (wild-type and B.1.1.7).**

Neutralization assays were conducted as previously described.^23^ Serial dilutions of plasma samples (heat inactivated at 56 °C for 30 min) were prepared with DMEM-C media and incubated with WT or B1.1.7 pseudotyped virus for 1 h at 37 °C in 96-well plates. Next, HeLa cells stably expressing the ACE2 receptor (provided by Dr James Voss, Scripps Research, La Jolla, CA) were added (12,500 cells/50µL per well) and the plates were left for 72 hours. Infection level was assessed in lysed cells with the Bright-Glo luciferase kit (Promega), using a Victor™ X3 multilabel reader (Perkin Elmer).  Measurements were performed in duplicate and the duplicates used to calculate the ID_50_ using GraphPad Prism.

**Interferon-gamma (IFNγ) and Interleukin-2 (IL-2) producer cell detection by Fluorospot**

Cryopreserved PBMC from vaccinated individuals were thawed and stimulated with peptide pools spanning the receptor binding domain (RBD) or the C-terminal S2 domain of the SARS-CoV-2 spike protein (S-2), CEF (CMV, EBV, influenza virus HLA class I epitopes) + CEFT (CMV, EBV, influenza virus, tetanus toxoid HLA class II epitopes) as a positive control (JPT Peptides, final concentration of 0.25µg/ml/peptide) or DMSO, as a negative control. IFNγ/IL-2 responses were detected by direct FluoroSpot assays (MabTech, Sweden) according to the manufacturer’s instructions. Briefly, after resting for 1 hour, 2x10^5^ PBMC were transferred to wells of a pre-coated FluoroSpot plate and stimulated in triplicate for 24h with the stimuli described above. Afterward, cells were removed by washing, and antibody-bound cytokine identified using a cocktail of anti-IL-2 and anti-IFN-γ detection antibodies followed by two fluorescent conjugates according to manufacturer’s instructions. Plates were scanned using AID *i*Spot Spectrum reader and analysed using AID EliSpot 8.0’ software (AID Autoimmun Diagnostika). Data were analysed using a QC standard operating procedure and values are expressed as cytokine secreting cells/10^6^ PBMC following subtraction of values from negative control wells. For every batch, an unvaccinated control was run as a negative control and a vaccinated individual with prior SARS-CoV-2 infection was included as a positive control.

**Supplementary Tables**

**Table 1: Routine full blood count performed at each trial specific visit.**

| Routine blood parameters | | | | |
| --- | --- | --- | --- | --- |
| Timepoint | **Pre-Vaccine** | **3 weeks following one dose of BNT162b1 vaccine** | **5 weeks following one dose of BNT162b1 vaccine** | **At 5 weeks following D1 & D21 dosing** |
| Solid Cancers | | | | |
| Total numbers | 75 | 65 | 21 | 19 |
| Hb | 121 (79-175) | 119 (15-152) | 113 (93-135) | 121 (81-149) |
| WCC | 6.5 (3-19) | 6.3 (2.1-25) | 4.3 (1.1-18) | 6.3 (3.8-11.6) |
| Platelets | 245 (90-688) | 231 (195-668) | 210 (52-989) | 276 (122-411) |
| Neutrophils | 4 (1.2-16) | 4.2 (1-22.5) | 2.6 (0.7-15.3) | 4.6 (1.9-7.8) |
| Lymphocytes | 1.5 (1.2-16) | 1.4 (0.4-5) | 1.1 (0.4-3.2) | 1.5 (0.7-4.7) |
| Basophils | 0.1 (0-1.5) | 0.1 (0-0.4) | 0.1 (0-0.2) | 0.1 (0-0.2) |
| Eosinophils | 0.1 (0-0.8) | 0.1 (0-1.9) | 0.2 (0-0.5) | 0.1 (0-0.2) |
| Monocytes | 0.8 (0-1.5) | 0.6 (0.1-1.9) | 0.7 (0-1.6) | 0.6 (0.3-1) |
| CRP | 14.2 (0-78) | 4 (1-149) | 3 (1-111) | 2 (1-45) |
| Haematological Cancers | | | | |
| Total numbers | 38 | 34 | 8 | 2 |
| Hb | 127 (70-127) | 119 (74-171) | 116.5 (95-146) | N/A |
| WCC | 5.2 (1-101) | 4.6 (0.5-51.7) | 3.6 (1.1-18) |  |
| Platelets | 185 (13-473) | 166 (51-514) | 145 (52-276) |  |
| Neutrophils | 5.5 (0.1-13.1) | 2.7 (0.4-22.9) | 2.1(0.7-15.3) |  |
| Lymphocytes | 1.6 (0.2-97) | 1.05 (0.1-179) | 0.8 (0.4-3.2) |  |
| Basophils | 0.1 (0-0.9) | 0.1 (0-0.3) | 0.1 (0-0.1) |  |
| Eosinophils | 0.1 (0-0.5) | 0.1 (0-0.7) | 0.2 (0-0.3) |  |
| Monocytes | 0.8 (0.1-2.5) | 0.5 (0-2.1) | 0.5 (0-1.6) |  |
| CRP | 2 (1-144) | 3 (1-75) | 1.5 (1-15) |  |

**Table 2: List of antibodies for the antibody staining mix for flow cytometry**

| **Company** | **Catalog number** | **Description** | **Format** | **Dilution** |
| --- | --- | --- | --- | --- |
| Biolegend | 318310 | CD56 | APC | 1:50 |
| Biolegend | 325610 | CD14 | AF488 | 1:100 |
| Biolegend | 304026 | CD45 | PerCP | 1:200 |
| Biolegend | 317342 | CD3 | APC-Cy7 | 1:200 |
| Biolegend | 302208 | CD19 | PE | 1:200 |
| Biolegend | 302016 | CD16 | PE-Cy7 | 1:800 |
| Biolgend | 323032 | CD15 | BV605 | 1:400 |
| Biolegend | 312226 | CD10 | BV711 | 1:40 |
| Biolegend | 305020 | CD64 | BV421 | 1:40 |
| Biolegend | 304830 | CD62L | BV785 | 1:100 |
| Biolegend | 302240 | CD19 | BV786 | 1:100 |
| Biolegend | 301830 | CD14 | BV421 | 1:100 |
| Biolegend | 354208 | CD303 | FITC | 1:100 |
| Biolegend | 305406 | CD86 | PE | 1:100 |
| BD | 563083 | HLA-DR | BV510 | 1:100 |
| Biolegend | 331524 | CD1c | APC | 1:100 |
| Biolegend | 301638 | CD11c | BV650 | 1:100 |
| Biolegend | 306016 | CD123 | PerCP-Cy5.5 | 1:100 |
| Biolegend | 317340 | CD3 | AF700 | 1:200 |
| Biolegend | 302016 | CD16 | PE-Cy7 | 1:200 |
